# Normative Executive Function Development Reveals Age-Varying Mental Health Associations in Youth

**DOI:** 10.1101/2025.09.04.25335085

**Authors:** Yang Li, Lirou Tan, Yinan Duan, Xiaoyu Xu, Haoshu Xu, Mei Yu, Luxia Jia, Zhilin Li, Chenguang Zhao, Qunlin Chen, Bart Larsen, Adam Pines, Tengfei Wang, Runsen Chen, Zaixu Cui

## Abstract

**IMPORTANCE:** Executive function (EF) is crucial for adolescent development and mental health. However, population-level benchmarks of EF growth and their relevance to psychiatric symptoms remain unclear, especially for non-Western populations.

**OBJECTIVE:** To establish normative developmental charts of EF across adolescence and examine how deviations from these norms relate to mental health symptoms in an age-specific manner.

**DESIGN:** Primary data were drawn from the baseline data of the ongoing Adolescent Health Enhancing Long-term Plan (A-HELP) study (2022-2027), with replication from three waves of longitudinal data (2016-2022) from the Adolescent Brain Cognitive Development (ABCD) study.

**SETTING:** Population-based studies conducted in China (A-HELP).

**PARTICIPANTS:** The A-HELP sample included 33,622 Chinese adolescents (11.00-18.00 years; 16,558 males) who completed EF tasks and mental health assessments. Normative developmental charts were constructed using generalized additive models for location, scale, and shape, from which individual EF deviation scores were derived. Associations with mental health were examined using generalized additive models, including age-varying interaction analyses. Replication were performed in 11,549 U.S. adolescents (8.92-15.75 years; 6,010 males) from the ABCD study.

**MAIN OUTCOMES AND MEASURES:** Three EF tasks were assessed: Go/No-Go (inhibitory control), 1-back and 2-back (working memory). Mental health symptoms were measured using the Strengths and Difficulties Questionnaire, including emotional symptoms, conduct problems, peer problems, hyperactivity/inattention, and prosocial behavior.

**RESULTS:** Adolescents completed Go/No-Go (N=17,021), 1-back (N=15,945), and 2-back (N=10,167) tasks. All tasks showed significant age-related improvement and decreasing inter-individual variability. Higher EF deviation scores, which reflect better performance, were associated with fewer peer and conduct problems, lower hyperactivity/inattention, and greater prosocial behavior. Age-resolved analyses revealed that these associations varied across development, with stronger effect sizes observed in early adolescence that declined by late adolescence. Findings were replicated in 22,831 Flanker task observations from the ABCD study, showing consistent developmental patterns and EF-mental health associations.

**CONCLUSIONS AND RELEVANCE:** This study establishes normative developmental charts of EF in adolescence and highlights that deviations from these norms are linked to psychiatric symptoms, especially in early adolescence. These findings provide a developmental framework for identifying youth at risk of mental health difficulties, offering culturally generalizable benchmarks for early screening and intervention.

## Introduction

Executive function (EF) encompasses a set of cognitive processes that enable individuals to regulate behavior, adapt flexibly to changing demands, and achieve goals^1,2^. Among its core components, working memory supports the maintenance and manipulation of information, while inhibitory control allows for the suppression of irrelevant or inappropriate responses^1^. EF undergoes prolonged development during adolescence, shaping lifelong cognitive and behavioral outcomes^3–5^. Perturbations in this developmental course have been linked to a wide range of psychiatric symptoms^6,7^, including the emotional and behavioral difficulties commonly observed in youth. Mapping the normative course of EF development is therefore critical for identifying early risk markers of mental health problems and guiding targeted interventions.

While EF development has been a central focus of adolescent cognitive neuroscience, most studies have relied on relatively small samples^8–10^. A recent large-scale study established a canonical trajectory of EF maturation in adolescence, providing a benchmark for normative development in Western youth^4^. However, comparable population-based EF trajectory remains scarce for non-Western youth. Moreover, few studies have examined whether deviations from these normative trajectories are informative for mental health outcomes, such as hyperactivity/inattention, internalizing, and externalizing problems.

While associations between cognition and psychopathology are well documented, these relationship are often modest in size^11,12^, partly reflecting the common practice of treating wide age ranges as homogeneous^13,14^. Such approaches risk obscuring heterogeneity in development, particularly during adolescence, a period marked by rapid and nonlinear neurocognitive change^4,15^. Addressing these gaps requires large, non-Western datasets capable of defining normative EF development and capturing its relevance to general mental health, within a developmentally sensitive framework.

To this end, we constructed normative developmental charts for working memory and inhibitory control using data from more than 33,000 Chinese adolescents aged 11 to 18. These charts showed both continuous improvements in EF performance and a decline in inter-individual variability, suggesting developmental convergence. Using these references, we derived individualized, age-normed EF deviation scores and examined their associations with psychiatric symptoms reported on the Strengths and Difficulties Questionnaire (SDQ), covering emotional, behavioral, and social domains. We found that higher EF deviation scores were associated with fewer peer problems and conduct problems, lower hyperactivity/inattention, and greater prosocial behavior, although the effect sizes were small. Age-resolved analyses further revealed that effect sizes varied across adolescence, peaking at early adolescence. Replication in the Adolescent Brain Cognitive Development (ABCD) study^16^, an independent U.S. cohort of over 10,000 youths, confirmed consistent EF trajectories, declining variability, and developmentally modulated associations with mental health problems. Together, these findings provide large-scale, culturally generalizable benchmarks for adolescent EF development and underscore the value of individualized, developmentally informed EF deviations for identifying youth at risk for mental health difficulties.

## Methods

### Study design and participants

Adolescents aged 11-18 were recruited from Yunfu City, Guangdong, China, as part of the Adolescent Health Enhancing Long-term Plan (A-HELP) cohort, a longitudinal project on youth mental health and cognition (**Fig. S1**). Surveys and cognition tests were conducted in classrooms across 259 schools, with appropriate consent and IRB approval. EF was assessed via inhibitory control (Go/No-Go) and working memory (1- and 2-back). Of 46,947 tested, participants with missing or inconsistent demographics, out-of-range age, ≤ 50 valid trials, accuracy ≤ 20%, or extreme performance were excluded (**Fig. S2**), yielding 33,622 adolescents (Go/No-Go: 17,021; 1-back: 15,945; 2-back: 10,167; **Table S1; Fig. S3A-C**). See **Supplementary Text** for details.

### Assessment of inhibitory control and working memory

Inhibitory control was assessed using the Go/No-Go task, requiring responses to frequent “Go” stimuli and withholding responses to infrequent “No-Go” stimuli (**Fig. S4A**). Performance was quantified using *d’* (Z(hit rate) – Z(false alarm rate)), with higher values indicating stronger inhibitory control. Working memory was measured using 1-back and 2-back tasks, where participants responded to stimulus repetitions one or two trials earlier (**Fig. S4 B & C**). Accuracy, the proportion of correct responses, served as the primary index of working memory. Full task details are provided in the **Supplementary Text**.

### Mental health assessment

Mental health was assessed using the adolescent self-report Strengths and Difficulties Questionnaire (SDQ), a validated 25-item measure of emotional and behavioral problems (Chinese version)^17^. Participants rated items on a 3-point scale (0-2) based on the past six months. The SDQ includes five subscales, each comprised of five items: emotional, peer, conduct, hyperactivity/inattention, and prosocial. Higher scores indicate greater difficulties, except prosocial, where higher scores reflect positive behavior. Each subscale was analyzed separately.

### Normative developmental charts with GAMLSS

Normative developmental charts for EF were constructed using the Generalized Additive Models for Location, Scale, and Shape (GAMLSS)^18,19^, implemented via the *gamlss* in R (v4.2.2). GAMLSS models multiple distributional moments, including mean (μ), variance (σ), skewness (ν), and kurtosis (τ), as smooth age-dependent functions, and therefore enables characterization of linear and nonlinear age-related changes in both central tendency and variability. For each EF measure, age-related effects on μ and σ were modeled using penalized B-splines^20^, sex was included as a fixed effect^20^, and ν and τ were set as constants. Distribution family and spline complexity were optimized via Bayesian Information Criterion (BIC) and model convergence. Using the full sample, we estimated developmental percentiles (5^th^-95^th^) and age-related variance trajectories. Model fit was assessed with quantile residual and 10,000 bootstrap iterations. See **Supplementary Text** for details.

### Individualized deviations from normative EF trajectories

To quantify deviations from normative EF development, we computed individualized deviation scores based on the GAMLSS-derived age- and sex-specific distributions. The full sample was stratified by sex and randomly split in half. Each subset was alternately used as training to fit the model and test set to compute deviations. For each test participant, GAMLSS parameters (μ, σ, ν, τ) defined the normative distribution at their age and sex. The observed EF score’s centile rank under this distribution was then transformed into a deviation z-score.

### Association between EF deviations and mental health

We assessed associations between EF deviation scores and SDQ subscales using generalized additive models (GAMs), controlling for age and sex. Effect sizes were quantified with correlation coefficients and standardized regression coefficients (β). Significance was evaluated by comparing full vs. reduced models using parametric bootstrap likelihood ratio statistic^21^, with Bonferroni correction for multiple comparisons.

To investigate whether these associations varied with age, we fitted GAMs with smooth interaction terms between age and EF deviation scores. Standardized slope were estimated at 1,000 evenly spaced age points between 11 and 18 years^22,23^. This enabled age-resolved mapping of how the strength and direction of the EF-mental health associations evolved over adolescence. At each age point, the estimated slope was compared to the whole-sample β coefficient to evaluate whether effect sizes were amplified at specific developmental stages.

### Replication in the ABCD study

We used longitudinal data from release 5.1 of the Adolescent Brain Cognitive Development (ABCD) study^16^ as replication. Inhibitory control was assessed using the Flanker task (**Fig. S4D**), with a composite of accuracy and reaction time to index performance. After exclusions, the sample included 11,549 participants at baseline (8.92-11.08 years), 8,068 at 2-year (10.58-13.83 years), and 3,214 at 4-year follow-up (12.50-15.75 years) (**Fig. S3D**). Working memory was not analyzed due to its absence at 2-year follow-up. Mental health was assessed using the Child Behavioral Checklist (CBCL)^24^, focusing on internalizing, externalizing, social, and attention problems, which align with SDQ dimensions of emotional, conduct, peer, and hyperactivity problems^25,26^. No CBCL domain corresponds to prosocial. Longitudinal EF-mental health associations were tested with mixed-effects models including participant random intercepts. CBCL z-scores were outcomes, EF deviation z-score predictors, with age and sex as covariates. Age-varying effects were assessed via interaction terms between age and EF deviation, and standardized slopes were estimated across 1,000 age points. Model details are in the **Supplementary Text**.

## Results

The final analytic sample included 33,622 participants (age: 11.00–18.00 years; 16,558 males) in total, with 17,021participants for the Go/No-Go task (8,485 males), 15,945 participants for the 1-back task (7,690 males), and 10,167 participants for the 2-back task (4,468 males) from A-HELP cohort (**Fig. S3** & **Table S1**).

### Normative developmental charts for executive function

Using GAMLSS, we estimated normative developmental charts for three EF tasks from 11 to 18. All tasks showed significant improvements across adolescence. The Go/No-Go task demonstrated a linear increase in *d’*, indicating steady maturation of inhibitory control without plateauing by age 18 (**Fig. 1A**, left). In contrast, both 1-back and 2-back tasks showed significant developmental gains in accuracy that plateaued around late adolescence (**Fig. 1B-C**, left). The 2-back task (**Fig. 1C**), which imposes greater demands on working memory than the 1-back (**Fig. 1B**), began with markedly lower accuracy in early adolescence but exhibited a steeper developmental trajectory. The growth rates of EF performance varied slightly across populations centiles, including the 5^th^, 25^th^, 50^th^ (median), 75^th^, and 95^th^ centiles (**Fig. 1 A–C**, left). This result suggests that adolescents with varying levels of task performance may show subtly different developmental trajectories. For all three tasks, inter-individual variability declined steadily with age, indicating convergence toward normative functioning (**Fig. 1A-C**, right). These patterns were confirmed by bootstrapped confidence intervals and first-derivative analyses marking age intervals of significant change (bars below scatter plots in **Fig. 1**).

**Fig. 1.**
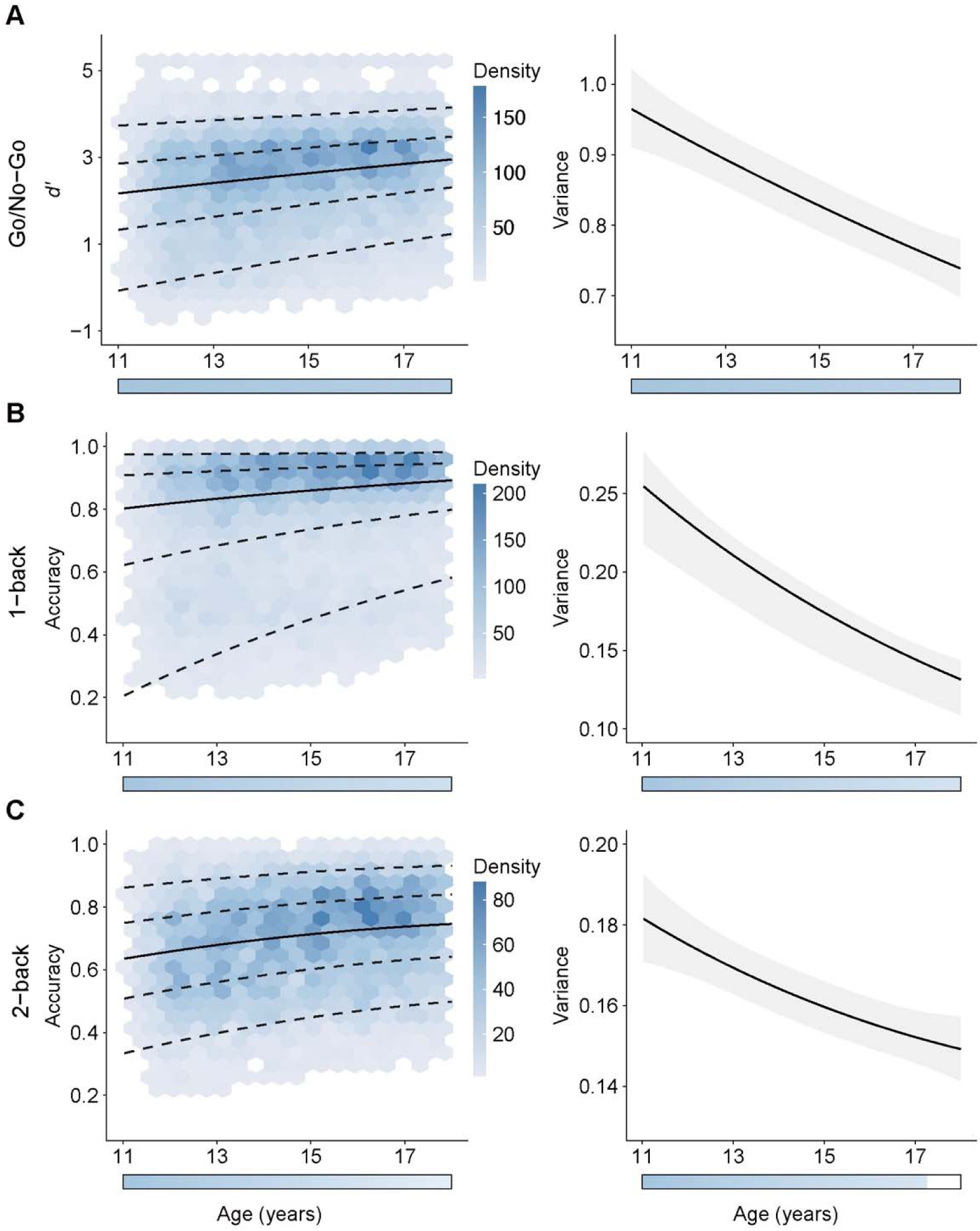
Normative developmental charts of executive function during adolescence. Age-related developmental trajectories for (**A**, left) Go/No-Go *d’* (inhibitory control), (**B**, left) 1-back accuracy, and (**C**, left) 2-back accuracy (working memory) in a large Chinese adolescent cohort (ages 11–18 years). Quantile curves were estimated using GAMLSS, modeling non-linear age effects on multiple distributional moments. Solid lines denote median performance; dashed lines represent the 5^th^, 25^th^, 75^th^, and 95^th^ percentiles. Hexagonal bins indicate participant density, with darker regions reflecting higher participant density. Age-related changes in inter-individual variability, measured as model-derived variation for (**A**, right) Go/No-Go, (**B**, right) 1-back, and (**C**, right) 2-back tasks. Solid lines indicate fitted variance trajectories; shaded areas show 95% confidence intervals derived from 10,000 bootstrap iterations. Horizontal color bars denote age periods with significant developmental changes, based on the first derivative of the fitted trajectory. GAMLSS: generalized additive models for location, scale, and shape.

### Associations between EF deviations and mental health

Using the normative developmental charts, we derived deviation z-scores for each adolescent’s EF performance relative to age norms. Higher deviation scores indicated better performance. We examined associations between EF deviations and mental health outcomes, measured by the SDQ, adjusting for age and sex (**Fig. 2**). Across all three EF tasks, higher EF deviations were significantly associated with fewer peer problems (Go/No-Go: *r* = −0.09, β = −0.09, *P_Bonf_* < 0.0015; 1-back: *r* = −0.11, β = −0.10, *P_Bonf_* < 0.0015; 2-back: *r* = −0.12, β = −0.10, *P_Bonf_* < 0.0015), and more prosocial behavior (Go/No-Go: *r* = 0.07, β = 0.08, *P_Bonf_* < 0.0015; 1-back: *r* = 0.07, β = 0.07, *P_Bonf_*< 0.0015; 2-back: *r* = 0.08, β = 0.08, *P_Bonf_* < 0.0015). Better Go/No-Go performance was also associated with lower hyperactivity symptoms (*r* = −0.05, β = −0.04, *P_Bonf_*< 0.0015). Better inhibitory control and low-load working memory were significantly associated with fewer conduct problems (Go/No-Go: *r* = −0.02, β = −0.03, *P_Bonf_* < 0.0105; 1-back: *r* = −0.02, β = −0.03, *P_Bonf_*< 0.0120). No significant associations were observed between EF deviations and emotional symptoms across all three tasks (Go/No-Go: *r* = 0.01, β = 0.01, *P_Bonf_* = 1.0000; 1-back: *r* = 0.02, β = 0.02, *P_Bonf_* = 0.6689; 2-back: *r* = 0.01, β = 6.8 × 10^−4^, *P_Bonf_* = 1.0000), between high-load working memory (2-back) deviations and conduct problems (*r* = −0.01, β = −0.03, *P_Bonf_*< 0.1605), or between working memory tasks and hyperactivity (1-back: *r* = −0.02, β = −0.02, *P_Bonf_*= 0.3990; 2-back: *r* = −0.03, β = −0.02, *P_Bonf_* < 0.1710). Together, these findings suggest that adolescents who perform better than age-expected norms in EF tasks tend to exhibit fewer behavioral problems and greater prosocial behaviors.

**Fig. 2.**
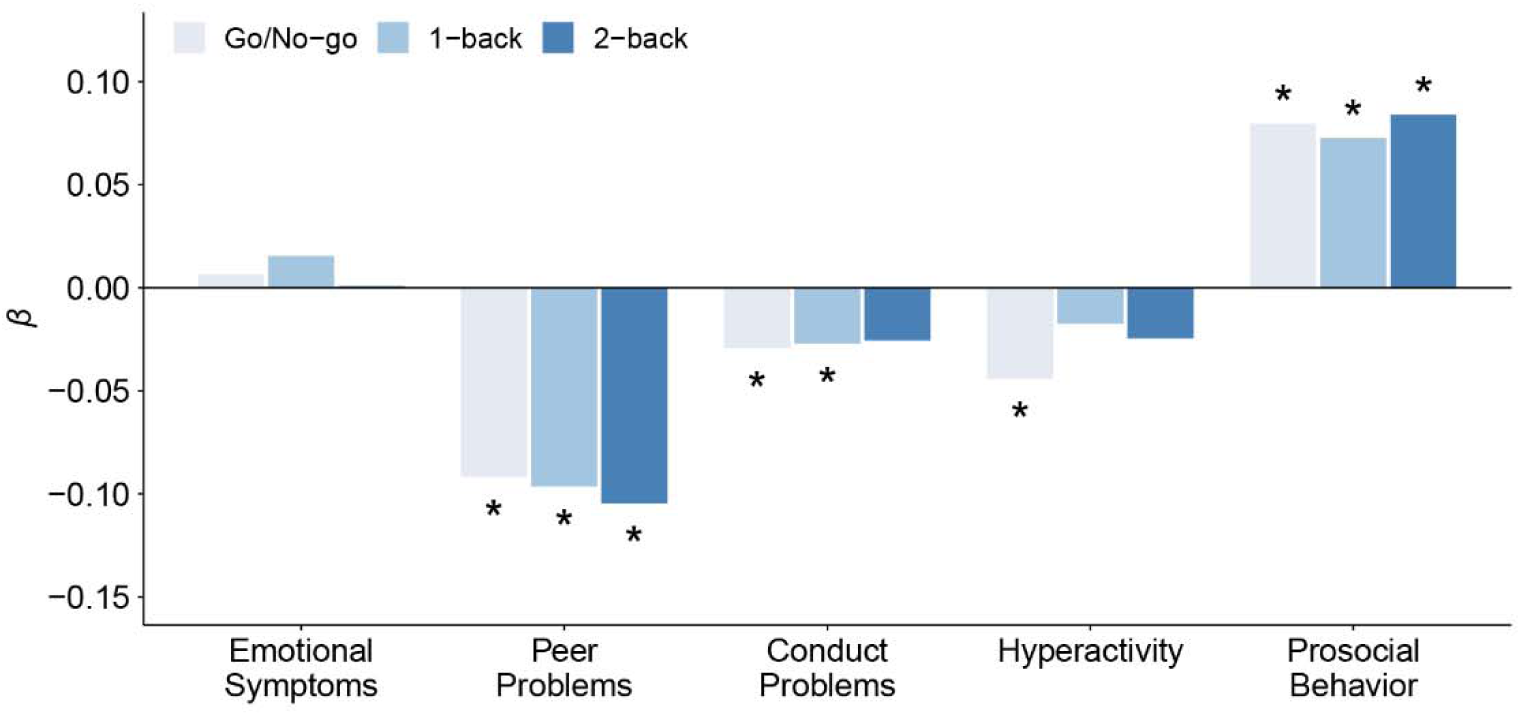
Associations between EF deviations and mental health dimensions. Standardized regression coefficients (β) represent associations between EF deviation z-scores and five SDQ subscales: emotional symptoms, peer problems, conduct problems, hyperactivity, and prosocial behavior. Results are shown for three executive function tasks: Go/No-Go (light blue), 1-back (medium blue), and 2-back (dark blue). All associations were estimated using GAMs controlling for age and sex. Asterisks indicate associations that survived Bonferroni correction at *P_Bonf_* < 0.05. Higher positive EF deviation z-scores indicate better performance relative to age-based normative expectations. EF: executive function. GAM: generalized additive model. SDQ: strengths and difficulties questionnaire.

### Age-varying associations between EF deviations and mental health

We next examined whether the associations between EF deviations and mental health varied with age by estimating age-specific slopes (i.e., standardized regression coefficients) across adolescence (**Fig. 3**). These analyses allowed us to identify developmental periods when associations were stronger or weaker than the full-sample estimates. Across nearly all EF tasks and SDQ dimensions, the estimated slopes varied with age and often diverged from the overall beta coefficients derived from the full sample (**Fig. 3**). In many cases, age-specific effect sizes were notably stronger during early adolescence (ages 11–13) and gradually weakened with age. For example, the association between 1-back deviation scores and hyperactivity symptoms peaked around age 11 with a slope nearing −0.13 (**Fig. 3**), whereas the overall association using the full sample was close to −0.02 (see **Fig. 2**). Similarly, the Go/No-Go task showed stronger negative associations with hyperactivity during early adolescence.

**Fig. 3.**
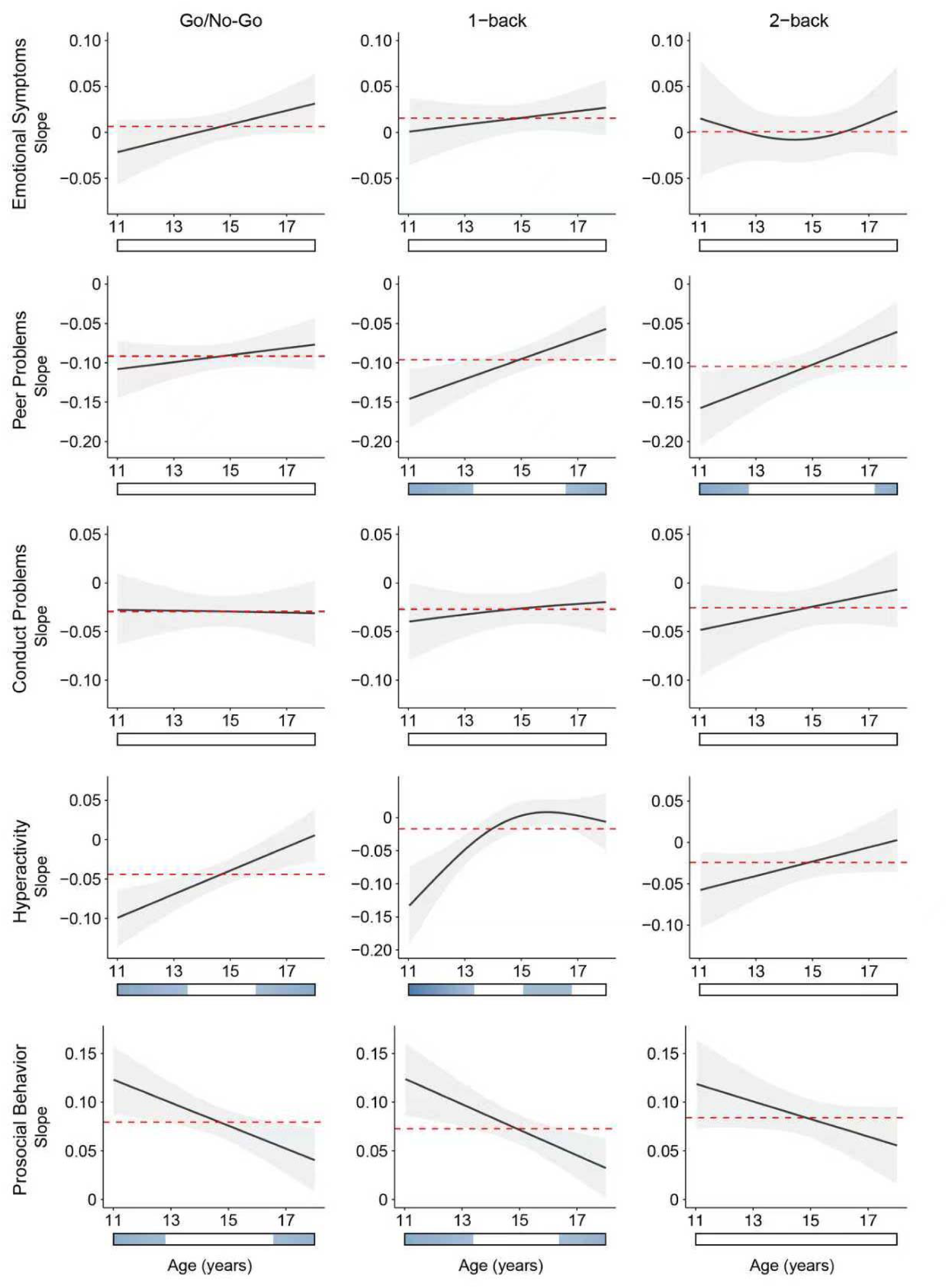
Age-resolved associations between EF deviation and mental health symptoms. Each panel depicts the age-varying standardized slope (y-axis) representing the association between EF deviation z-scores and SDQ subscales across adolescence (x-axis, ages 11–18 years). Results are shown separately for the three EF tasks: Go/No-Go (left), 1-back (middle), and 2-back (right). Shaded areas represent 95% confidence intervals from the GAM-based interaction models. More negative slopes indicate stronger negative associations between better EF and more severe symptoms, while positive slopes reflect positive associations. Red dotted horizontal lines mark the beta coefficients from models using the full sample, as reported in **Fig. 2**. Higher EF deviation scores relative to age norms indicates better performance. EF: executive function. SDQ: strengths and difficulties questionnaire.

Comparable developmental patterns were observed for peer problems and prosocial behavior: better EF performance was most strongly associated with reduced peer problems and increased prosocial behavior in early adolescence than in later years. Notably, 1-back and 2-back deviation scores showed more negative slopes for peer problems during this period, indicating greater protective effects of higher EF. For prosocial behavior, Go/No-Go and 1-back deviation scores exhibited stronger positive associations at younger ages. In contrast, emotional symptoms and conduct problems showed relatively weak and age-invariant slopes, aligning with the small associations observed in the full-sample.

Overall, these findings reveal that the strength of associations between EF deviations and mental health symptoms is not uniform across adolescence. Instead, many relationships appear strongest earlier in development, particularly around ages 11 to 13, and attenuate by late adolescence. This suggests that executive function may play a more prominent role in shaping mental health outcomes during earlier developmental windows.

### Replication in the ABCD dataset

We constructed normative growth charts for Flanker performance (**Fig. 4A**) and demonstrated the developmental decline of inter-individual variability (**Fig. 4B**) using ABCD dataset, replicating the pattern observed in the A-HELP cohort. Higher EF deviations were significantly associated with fewer social problems (*r* = −0.03, β = −0.04, *P_Bonf_* < 0.0004), externalizing symptoms (*r* = −0.01, β = −0.03, *P_Bonf_* < 0.0004), and attention problems (*r* = −0.04, β = −0.04, *P_Bonf_* < 0.0004) (**Fig. 4C**). The association with internalizing symptoms was not significant (β = −4.27 × 10^3^, *P_Bonf_* = 1.0000). These findings were consistent with A-HELP data. Finally, we replicated the development of age-specific slopes. In A-HELP, age-resolved associations were observed for inhibitory control (Go/No-Go), specifically with hyperactivity/inattention and prosocial behavior (**Fig. 3**). Guided by these results, we focused replication in ABCD on CBCL attention problems (no CBCL counterpart exists for prosocial). We found that negative slopes between EF deviations and attention problems were stronger in early adolescence than in the full sample, paralleling A-HELP findings (**Fig. 4D**). Together, these replications support the generalizability of normative EF deviation as a developmentally dynamic marker of adolescent mental health.

**Fig. 4.**
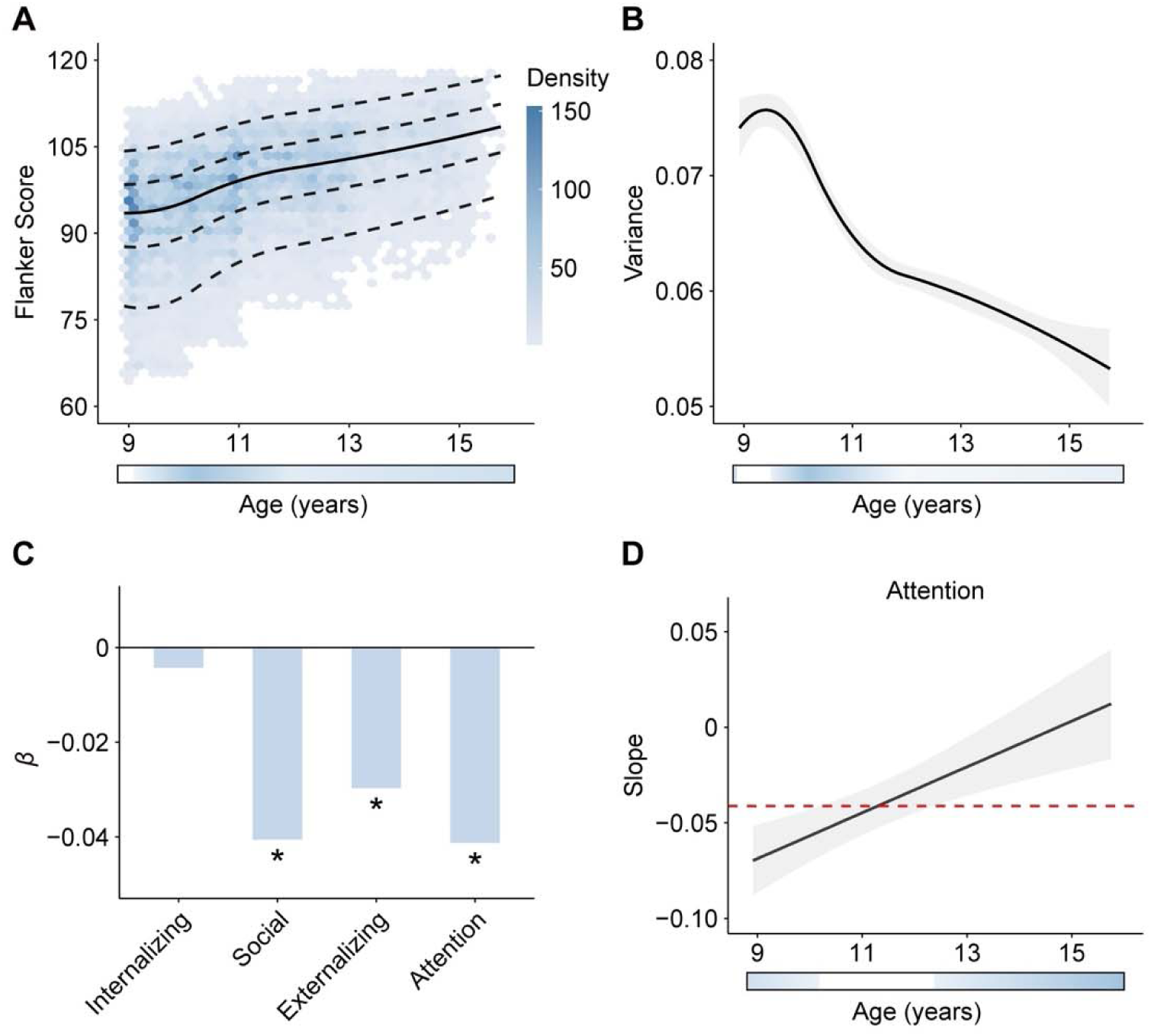
Replication of normative EF development and its association with mental health in the ABCD dataset. **A**, Normative developmental trajectories for inhibitory control measured by Flanker task performance in children aged 8.92–15.75 years. Curves depict the median (solid line) and 5^th^, 25^th^, 75^th^, and 95^th^ percentiles (dashed lines) estimated using GAMLSS. Density shading represents participant distribution across age. **B**, Age-related decline in inter-individual variability, quantified by the modeled variance and 95% bootstrap confidence intervals. **C**, Associations between Flanker EF deviation z-scores and four mental health dimensions from the CBCL: internalizing, externalizing, attention and social problems. Bars show standardized β coefficients; asterisks denote Bonferroni-significant associations (*P_Bonf_* < 0.05.). **D**, Age-resolved associations between EF deviation and CBCL attention symptom. Plots show the estimated slope of the relationship across age, with shaded areas indicating 95% confidence intervals from GAMMs. Higher EF deviation scores relative to age norms indicates better performance. EF: executive function. CBCL: Child Behavioral Checklist. GAMLSS: generalized additive models for location, scale, and shape.

## Discussion

In a large, population-based cohort of Chinese adolescents, with replication in a U.S. sample, we observed marked improvements in EF performance from early to late adolescence, accompanied by declining inter-individual variability. Adolescents performing above age-specific EF norms showed fewer peer problems and conduct problems, lower hyperactivity, and greater prosocial behavior. Notably, these associations varied by age, with the strongest effect sizes emerging in early adolescence. Together, these findings offer a developmentally anchored framework for quantifying cognitive deviations relevant to youth mental health.

Across ages 11 to 18 years, inhibitory control, measured with the Go/No-Go task, and working memory, assessed via the 1-back and 2-back tasks, both showed significant age-related gains, alongside reduced variability between individuals. These normative trajectories suggest that adolescents become progressively more capable of suppressing prepotent responses and managing interference^27^, while concurrently improving the capacity to maintain, update, and manipulate information^28^. The observed decline in variability may reflect a developmental convergence toward more stable, adult-like cognitive function, consistent with recent reports of reduced variability in brain connections associated with higher-order cognition during adolescence^29^. By establishing large-scale normative EF charts in more than 33,000 Chinese adolescents, this work addresses a critical gap, offering culturally relevant benchmarks that complement existing Western-based references^30^.

Working memory performance peaked near age 18, whereas inhibitory control continued to improve across the examined age range. This temporal disassociation aligns with prior findings in Western cohorts, where N-back performance plateaus around ages 18-20, whereas Stroop performance improves into adulthood, as observed in the Philadelphia Neurodevelopmental Cohort^30^. Moreover, the 2-back task, which imposes greater demands than the 1-back, began with lower performance and exhibited a steeper developmental trajectory. This pattern suggests that the more demanding aspects of working memory, such as continuous updating, sustained maintenance, and temporal order tracking^28^, undergo more pronounced maturation during adolescence.

Deviations from normative EF development were consistently associated with multiple domains of mental health. Adolescents exceeding age-specific EF expectations tended to have fewer peer difficulties and conduct problems, lower hyperactivity, and greater prosocial behavior. These findings align with evidence that EF impairments constitute a transdiagnostic feature of psychiatric conditions, often emerging early in development^6,13^. Although the effect sizes were small, this is consistent with broader literature indicating that cognitive predictors of mental health typically yield small effects at the population level—effects that, while modest, are more likely to replicate across independent cohorts^31–33^. Crucially, our analyses revealed that these associations were not constant across development, challenging the implicit assumption in much of the prior literature that cognition-symptom relationships are linear across age^31,33^.

Age-resolved models revealed pronounced developmental modulation. For example, the overall linear association between 1-back performance and hyperactivity was negligible (β ≈ −0.02), yet age-specific analyses showed substantially stronger associations in early adolescence, peaking at β ≈ −0.13 at age 11. Similar age-dependent effects were observed for Go/No-Go performance and hyperactivity. These findings are consistent with ADHD research indicating that executive function deficits covaried with hyperactive/impulsive symptoms during childhood and early-adolescence^34^. Comparable patterns also emerged between EF task performance and peer problems as well as prosocial behavior. These findings extend prior work^31^ showing that such associations depend on symptom severity by identifying age as an additional, critical moderator. Future research should explicitly model such non-linearity, using flexible, age-resolved approaches to identify developmental periods when cognitive markers have the greatest predictive value for mental health outcomes.

Several limitations should be noted. First, the cross-sectional design and the observational nature of the A-HELP cohort limits causal inference, though findings were replicated in longitudinal ABCD dataset. Second, the age range of 11 to 18 years precludes characterization of earlier developmental surges in EF during childhood or stabilization in early adulthood. Future studies spanning broader developmental windows is warranted. Third, mental health symptoms were assessed through self- or caregiver-reported questionnaires, which may introduce reporting bias. Future research could incorporate multi-informant reports or employ clinician-administered interviews.

## Conclusions

This study establishes large-scale, culturally generalizable normative charts of EF development in adolescence and demonstrates age-dependent associations between deviations from these norms and psychiatric symptoms, with the strongest relationships in early adolescence. These findings highlight the importance of incorporating developmental stage into the assessment of cognitive functioning and underscore the potential value of age-specific EF measures for identifying youth at risk for mental health difficulties.

## Acknowledgments

This work is supported by the STI 2030-Major Projects (2022ZD0211300), Chinese Academy of Medical Sciences Innovation Fund for Medical Sciences (2024-I2M-ZD-013), the National Natural Science Foundation of China (82472059), Non-profit Central Research Institute Fund of Chinese Academy of Medical Sciences (2024-RC416-02), and Chinese Institute for Brain Research, Beijing (CIBR) funds.

## Author contributions

Z.C. and R.C. designed and supervised the study. R.C. and Z.C. led data acquisition for A-HELP. M.Y., L.J., X.X., and Z.L. designed the tasks and contributed to data acquisition. Y.L., L.T. and X.X. cleaned the cognitive data, and R.C. and Y.D. cleaned the clinical data. Y.L., L.T. and X.X. performed the data analyses. Z.L., H.X., C.Z., Q.C., B.L., A.P., and T.W. provided analytical support. Y.L., Z.C., L.T., and X.X. wrote the manuscript. All authors reviewed and edited the manuscript.

## Supplemental information

Supplementary Text

Fig. S1 to S9

Tables S1 to S3

## Data availability

The ABCD 5.1 data release used in this study was accessed via the NDA under DOI: 10.15154/z563-zd24 (https://nda.nih.gov/abcd). Fast-track data from the ABCD Study are also available through the NDA. Derived data from the A-HELP cohort will be made available upon reasonable request to the corresponding author.

## Code availability

All analyses codes are available at: https://github.com/CuiLabCIBR/Adolescent_EF_Mental_Health.

