## Supplementary Materials for "Normative Executive Function Development Reveals Age-Varying Mental Health Associations in Youth"

**Authors:** Yang Li^1,2#^, PHD; Lirou Tan^1,2,4#^, MS; Yinan Duan^3#^, MEd; Xiaoyu Xu^1,2,5#^, MM; Haoshu Xu^1,2,6^, BEng; Mei Yu^3^, PHD; Luxia Jia^3^, PHD; Zhilin Li^1,2^, PHD; Chenguang Zhao^1,2^, PHD; Qunlin Chen^7^, PHD; Bart Larsen^8^, PHD; Adam Pines^9^, PHD; Tengfei Wang^10^, PHD; Runsen Chen^3*^, PHD; Zaixu Cui^1,2*^, PHD

^#^These authors contributed equally to this work

**Supplemental information**

Supplementary Methods

Fig. S1 to S9

Tables S1 to S3

### **Supplementary Methods**

#### **Participants**

Adolescents were recruited from Yunfu City, Guangdong Province, China, as part of the Adolescent Health Enhancing Long-term Plan (A-HELP) cohort. A-HELP is an ongoing longitudinal project designed to evaluate mental health and cognitive development in adolescence and to provide tailored psychological interventions (**Fig. S1**). Baseline data were collected in 2023, with annual follow-ups in the next two years. Cognitive assessments and mental health surveys were administered in classrooms on a class-by-class basis across 259 schools, selected according to the availability of computer facilities and teacher participation. Written informed consent was obtained from participants aged 18 years or older, and assent was obtained from the guardians and participants under 18 years. The study was approved by the Institutional Review Board at Tsinghua University.

A total of 46,947 participants completed at least one executive function task and provided demographic and mental health information, including biological sex, date of birth, national identification number, and responses to standardized mental health questionnaires. Due to time constraints in school-based testing, most participants were randomly assigned to complete either the Go/No-Go task or the N-back task, which included both 1-back and 2-back conditions. Specifically, 23,606 participants completed the Go/No-Go task, and 23,949 completed the N-back task, with 23,936 completing the 1-back condition and 20,403 completing the 2-back condition. A small subset completed multiple tasks: 608 participants completed both the Go/No-Go and 1-back tasks, 587 completed both the Go/No-Go and 2-back tasks, and 587 participants completed all three conditions.

Rigorous quality control procedures were applied. Participants were excluded if age information was missing (Go/No-Go: 167; 1-back: 261; 2-back: 215); if age calculated as the difference between the testing date (i.e., the date of task completion) and date of birth recorded on the national identification number differed by more than one year from the self-reported age (Go/No-Go: 157; 1-back: 167; 2-back: 134); or if there was a mismatch between self-reported sex and sex recorded on the national identification number (Go/No-Go: 16; 1-back: 32; 2-back: 26). Those outside the target age range of 11-18 years were further excluded due to very small sample sizes in other age groups (Go/No-Go: 1,352; 1-back: 1,458; 2-back: 1,254). An illustrative example of the age distribution (9-25 years) confirmed that only a small proportion of participants fell outside the target range of 11-18 years (**Fig. S2**).

For trial-level quality control, responses with reaction times below 200 ms were discarded^1^, and participants with 50% or more of trials removed under this threshold were excluded from further analysis (Go/No-Go: 1,281; 1-back: 2,809; 2-back: 5,180). Task accuracy was calculated separately for the two conditions of each task: target and non-target conditions for the 1-back and 2-back tasks, and Go and No-Go conditions for the Go/No-Go task. Accuracy-based exclusions were applied independently for each task. Participants were excluded from the Go/No-Go, 1-back, or 2-back analyses if their accuracy was ≤ 20% in either condition of that task (Go/No-Go: 3,104; 1-back: 3,186; 2-back: 3,401). Subsequently, participants were excluded if their performance was outside the three standard deviations from the age-specific group mean (one-year bins); for the Go/No-Go task, this criterion was applied to both accuracy and *d’* values (Go/No-Go: 508; 1-back: 78; 2-back: 26).

Following all quality control procedures, the final sample comprised 17,021 participants (8,485 males) for the Go/No-Go task, 15,945 (7,690 males) for the 1-back task, and 10,167 (4,468 males) for the 2-back task. Detailed inclusion and exclusion criteria are shown in **Fig. S3**, and sample demographics are provided in **Table S1**.

#### **Assessment of inhibitory control and working memory**

Executive function was assessed using two well-established computerized paradigms with alphabetical stimuli: a Go/No-Go task to measure inhibitory control and an N-back task to measure working memory.

In the Go/No-Go task, each trial began with a fixation cross from 700 to 1,000 milliseconds, followed by a stimulus displayed for 1,000 milliseconds. The stimuli consisted of uppercase letters (A, B, J, L, R, S, T, X, Y, Z). All letters except “X” served as “Go” stimuli, requiring participants to press the space key as quickly and accurately as possible. The letter “X” served as the “No-Go” stimulus, to which participants were instructed to withhold any response. A total of 76 Go trials and 24 No-Go trials were included. Task performance was quantified using the signal detection index *d’* (calculated as Z(hit rate) – Z(false alarm rate)), where the hit rate was defined as the proportion of correct responses to Go stimuli and the false alarm rate as the proportion of incorrect responses to No-Go stimuli. This metric was selected because it provides a bias-corrected estimate of discriminative ability, independent of response bias, and has been widely adopted in the cognitive neuroscience literatures to sensitively capture individual differences in inhibitory control^2^. Higher *d’* scores indicate more effective suppression of prepotent responses and greater accuracy in discriminating between target and non-target stimuli^3,4^.

In the N-back task, each trial began with a central fixation cross presented for 500 milliseconds, followed by a stimulus displayed for 2,000 milliseconds. The stimuli consisted of uppercase letters (A, D, E, H, I, N, R, S, T, U). Participants were required to determine whether the current letter matched the one presented *N* positions earlier in the sequence, with target trials defined as matches and non-target trials as non-matches. Two levels of working memory load were implemented. In the 1-back condition, participants judged whether the current letter matched the one presented immediately before (21 target and 37 non-target trials). In the 2-back condition, participants judged whether the current letter matched the one presented two trials before (19 target and 37 non-target trials). Participants pressed the “J” key for matches and the “F” key for non-matches. Accuracy, defined as the percentage of correct responses, was chosen as the primary performance metric because it directly indexes the ability to correctly identify and recall target stimuli over short time intervals. Including both 1-back and 2-back conditions allowed for the dissociation of relatively simple maintenance processes from more complex updating operations.

Schematics illustrating the structure of both the Go/No-Go and N-back tasks are provided in **Fig. S4**.

##### Assessment of mental health symptoms

Mental health was assessed using the Strengths and Difficulties Questionnaire (SDQ)^5^, a widely validated behavioral screening tool for children and adolescents that evaluates emotional and behavioral functioning across both strengths and difficulties. The SDQ consists of 25 items, each rated on a 3-point Likert scale (0 = Not True, 1 = Somewhat True, 2 = Certainly True), referencing the participant’s behavior over the past six months. Items are organized into five subscales of five items each: (1) Emotional problems: frequent complains of headaches/stomachaches/sickness, often unhappy or downhearted, many worries, nervous or clingy in new situations, and many fears or easily scared; (2) Conduct problems: often has temper tantrums, generally disobedient to adults, fights with or bullies other children, often lies or cheats, and steals from home, school, or elsewhere; (3) Hyperactivity/inattention: restless or overactive, constantly fidgeting or squirming, easily distracted, thinks before acting (reverse-coded), and sees tasks through to the end (reverse-coded); (4) Peer problems: rather solitary, has at least one good friend (reverse-coded), generally liked by other children (reverse-coded), picked on or bullied, and gets on better with adults than peers; and (5) Prosocial behavior: considerate of others’ feelings, shares readily with others, helpful if someone is hurt/upset/ill, kind to younger children, and often volunteers to help others. Subscale scores were calculated by summing responses for each domain (range, 0-10), with higher scores on the first four subscales indicating greater difficulties and higher scores on the prosocial subscale indicating greater strengths. The SDQ has demonstrated adequate internal consistency and strong predictive validity for mental health outcomes in adolescent populations across diverse cultural contexts. A validated Chinese version of the SDQ was administered.

##### Normative charts of EF development

Normative developmental charts for each EF measure were constructed using the Generalized Additive Models for Location, Scale, and Shape (GAMLSS) framework via the *gamlss* package in R (version 4.2.2)^6,7^. Unlike conventional additive models that capture only non-linear changes in the mean, GAMLSS simultaneously models multiple distributional parameters, including the mean (*µ*), variance (*σ*), skewness (*ν*), and kurtosis (*τ*), as functions of explanatory variables such as age and sex. This capability enables a comprehensive characterization of developmental trajectories, capturing not only central tendencies but also age-related changes in variability. This approach has been recommended by the World Health Organization for constructing reference growth charts^6^. Model fitting proceeded in three stages: (1) selection of the optimal response distribution and link functions; (2) estimation of model parameters; and (3) evaluation of model fit and reliability.

###### (i) Selection of optimal distribution and link functions

To identify the best distribution family, we systematically evaluated 24 continuous candidate distribution families with three or more parameters to identify the best-fitting distribution for each EF measure. Notably, although the GAMLSS framework provides over 100 distributions, we restricted our search to 24 continuous families with three or more parameters. These families are recommended for modeling developmental and growth-related data, as they allow age-related variation not only in the mean and variance but also in skewness and kurtosis^8^. In each model, the EF score served as the dependent variable, while age (modeled as a smooth B-spline function) and sex were included as predictors of the mean (*μ*) and variance (*σ*) parameters. The skewness (*ν*) and kurtosis (*τ*) parameters were estimated as intercepts only. Model estimation was carried out using maximum likelihood, with convergence defined as changes in log-likelihood < 0.001 across successive iterations, with a maximum of 200 iterations. Optimal distribution families were selected based on the Bayesian Information Criterion (BIC) and model convergence. For the Go/No-Go and 2-back tasks, the skew exponential power type 3 (SEP3) distribution provided the best fit, while the SEP type 2 (SEP2) distribution was optimal for the 1-back task (**Fig. S5**). To model non-linear age effects on the *μ* and *σ*, we used B-spline functions with degrees of freedom ranging from 2 to 6 and polynomial degrees of 2 or 3. The best-fitting B-spline complexity was again selected based on BIC and model convergence, with all the three tasks optimally fit by df = 2 and polynomial degree = 2 (**Fig. S6**).

###### (ii) Normative model fitting

Following identification of the optimal distribution families and spline complexity, we fit the final GAMLSS models to the full sample for each EF measure. The GAMLSS framework allows each distributional parameter to be flexibly modeled as a function of explanatory variables. In our models, both the location (*μ*) and scale (*σ*) parameters were modeled as functions of age and sex, with age entered as a smooth term using B-spline transformations and sex as a fixed effect. The skewness (*ν*) and kurtosis (*τ*) parameters were treated as intercept-only terms, give the absence of clear developmental hypotheses or strong prior evidence supporting covariate dependence for these higher-order moments. Formally, the GAMLSS models for each EF outcome took the form:

*Y ~ D (μ, σ, ν, τ)*

where *D* denotes the selected distributions, that the SEP3 for Go/No-Go and 2-back, and SEP2 for 1-back, and each parameter was expressed as:

$$\text{g}_{\text{1}}\left( \text{μ} \right)\text{=}\text{β}_{\text{μ}\text{0}}\text{+}\text{bs}_{\text{μ}}\left( \text{age} \right)\text{+}\text{β}_{\text{μ}\text{1}}\left( \text{Sex} \right)$$

$$\text{g}_{\text{2}}\left( \text{σ} \right)\text{=}\text{β}_{\text{σ}\text{0}}\text{+}\text{bs}_{\text{σ}}\left( \text{age} \right)\text{+}\text{β}_{\text{σ}\text{1}}\left( \text{Sex} \right)$$

$$\text{g}_{\text{3}}\left( \text{ν} \right)\text{=}\text{β}_{\text{ν}\text{0}}$$

$$\text{g}_{\text{4}}\left( \text{τ} \right)\text{=}\text{β}_{\text{τ}\text{0}}$$

Here, g_k_() represents the default link functions specified by the chosen distribution family, $\text{β}$ terms denote intercepts or regression coefficients, and $\text{bs}$() denotes spline transformation.

Using the fitted models, we generated developmental trajectories of the 5^th^, 25^th^, 50th, 75^th^, and 95^th^ percentiles of each EF measure, as well as the estimated variance across the age span. These percentile curves provide normative reference charts against which individual performance can be compared.

###### (iii) Model validation and diagnostic assessment

We employed a combination of diagnostic plots and bootstrap resampling to assess the goodness-of-fit and reliability of the GAMLSS-derived developmental trajectories. To evaluate the model fit, we visually inspected the residuals using three standard diagnostic tools: the normalized quantile residual distributions^9^, quantile-quantile (Q-Q) plots, and detrended transformed Owen’s plots^10^. The normalized quantile residuals for all four EF measures approximated a normal distribution, as indicated by skewness and kurtosis values near zero and Filliben correlation coefficients approaching 1 (**Table S3**). The Q-Q plots showed that the residuals closely followed the diagonal reference line, with slopes near unity, further supporting the assumption of approximate normality (**Fig. S7**). Additionally, the detrended transformed Owen’s plots revealed that the majority of the nonparametric 95% confidence bands crossed the zero line (**Fig. S8**), suggesting a normal distribution of the residuals. Together, these diagnostic indicators support that the fitted GAMLSS models captured the empirical distributions of the EF data appropriately.

To assess model reliability and quantify uncertainty around the estimated trajectories, we conducted a nonparametric bootstrap procedure. For each EF measure, we performed 10,000 iterations of stratified resampling with replacement, preserving the original sex ratio in each resampled dataset. On each bootstrap sample, we re-estimated the full GAMLSS model and extracted predicted values for (i) the median developmental trajectories, (ii) its first derivative (i.e., rate of change), (iii) the developmental trajectory of the variance, and (iv) its first derivative. The 95% confidence intervals (CIs) for these predicted values were then computed across iterations to reflect model uncertainty. The resulting CIs for the median developmental trajectories were consistently narrow across the age range, indicating robust and reliable estimation of developmental patterns in both central tendency and variability (**Fig. S9**). Variance trajectories were derived from the predicted *σ* parameter with 95% CIs quantifying the reliability of age-related changes in inter-individual variability (**Fig. 1 A–C,** right). Using the finite-difference method, we then computed the first derivatives of both the median and variance trajectories and obtained pointwise 95% CIs across bootstrap replicates. Age intervals where the 95% CIs of the first derivative were significantly different from zero were interpreted as periods of significant developmental change, with the median derivatives corresponding to the bars on the left of **Fig. 1A–C** (age-specific rates of EF growth) and the variance derivatives corresponding to the bars on the right of **Fig. 1 A–C** (age-specific changes in inter-individual variability).

###### Individual deviation score computation

To quantify each participant’s deviation from the age- and sex-adjusted normative distribution, we employed a stratified cross-validation framework using the entire sample for each EF measure. Specifically, the full dataset was randomly split into two equal-sized subsets, ensuring identical sex distributions across subsets via stratified sampling. Each subset served alternately as the training and test set, enabling normative estimation and deviation scoring in a held-out manner.

Within each fold, we first fitted the normative GAMLSS model to the training subset using the same distributional family, B-spline complexity, and covariates (age and sex) established in the primary model fitting. These fitted models served as the normative reference for computing centile positions of individuals in the held-out test set.

For each test subject, we applied the estimated cumulative distribution function (CDF) of the trained GAMLSS model to compute their quantile position (i.e., the proportion of the normative distribution lying below their observed EF score, conditional on their age and sex). These quantile scores were then transformed into deviation z-scores by applying the probit (inverse-normal) function, thus yielding standardized deviation scores that reflect how far each individual departs from the age- and sex-expected norm in units of standard deviation.

This cross-validation approach preserves independence between model training and deviation scoring, avoiding overfitting and ensuring that individual deviations are assessed relative to normative expectations not influenced by the individual’s own data. The resulting z-scores were used in subsequent analyses examining associations between EF development and mental health outcomes.

##### Association between EF deviation and mental health

The relationships between EF deviation z-scores and mental health measurements were assessed using generalized additive models (GAMs) via the *mgcv* package in R^11^. For each of the five mental health subscales derived from the Strengths and Difficulties Questionnaire (SDQ), raw scores were standardized (z-scored) to facilitate interpretability and comparability across subscales. Extreme values beyond ±3 standard deviations were excluded.

The SDQ z-scores were regressed on EF deviation z-scores, controlling for age and sex. EF deviation z-scores served as predictors. Age was modeled using penalized thin plate regression splines with a maximum basis dimension (*k*) of 3 to reduce overfitting risk while allowing sufficient flexibility to model non-linear age effects. Sex was included as a categorical covariate. The regression model is summarized as:

SDQ z-scores ~ EF deviation z-scores + *s*(Age, *k* = 3, *fx* = FALSE) + Sex.

Model significance was evaluated by comparing full models (including EF deviation terms) against reduced null models (excluding EF deviation terms) using a parametric bootstrap method with 1,000 iterations based on the likelihood ratio test statistic^8,12,13^. Bonferroni correction was applied to control for multiple comparisons across EF subdomains and SDQ subscales. We additionally computed partial correlation coefficients (adjusted for age and sex) to quantify effect sizes.

To examine whether the strength of the associations between EF and mental health varied across age, we fit age-varying coefficient models. Specifically, GAMs were used to model the SDQ z-scores as a smooth function of age, stratified by levels of EF deviation z-scores. This approach allowed the age-dependent contribution of EF deviations to mental health to vary across the age span. The model structure is as follows:

SDQ z-scores ~ *s*(Age, by = EF deviation z-scores, *k* = 3, *fx* = FALSE) +

*s*(Age, *k* = 3, *fx* = FALSE) + Sex.

This model enables visualization and statistical inference regarding whether EF deviations show stronger or weaker associations with mental health at specific developmental windows, such as early versus late adolescence. To determine the age-specific relationship between EF deviation and SDQ scores, we drew 10,000 samples from the multivariate normal distribution defined by the estimated GAM coefficients and their covariance matrix. For each draw, we calculated the slope at 1,000 evenly spaced age points across the entire age span, where the slope was defined as the difference in fitted SDQ scores between the 90th and 10th percentiles of EF deviation values, divided by the corresponding difference in EF deviation values. This procedure produced a bootstrap distribution of age-specific slopes, reflecting the uncertainty in the EF–mental health association across development^14^. As shown in **Fig. 3**, at each age point, the black solid line represents the median of the posterior slope distribution, while the gray shaded area shows the 95% CI. The red dashed line denotes the overall linear effect (*β* coefficient) estimated from conventional regression models without age-varying terms. Age points where the overall *β* coefficient fell outside the 95% CI of the posterior slope distribution were considered to show significantly stronger or weaker EF effects compared to the overall association. These significant age intervals were further highlighted by bars below the main plot, with the color intensity of each bar proportional to the absolute difference between the age-specific slope and the overall *β* coefficient.

##### Replication in the ABCD study

To validate the robustness and generalizability of our findings, we conducted a replication analysis using the Adolescent Brain Cognitive Development (ABCD) Study Release 5.1. This independent cohort included 11,715 participants with three longitudinal acquisitions, including baseline (N = 11,715, 8.92-11.08 years), 2-year follow-up (N = 8,185, 10.58-13.83 years), and 4-year follow-up (N = 3,268, 12.50-15.75 years). Participants with missing sex information or outlier performance scores—defined as ±3 standard deviations from the age-specific mean within one-year bins—were excluded (N_baseline_ = 162; N_2-year_ = 104; N_4-year_ = 31), resulting in a final analytic sample of N_baseline_ = 11,549; N_2-year_ = 8,068; N_4-year_ = 3,214 (**Fig. S3D**).

In the ABCD study, only the Flanker task was administered consistently across all three time points to assess executive function. Working memory tasks were not included in the 2-year follow-up. Therefore, our replication analyses focused on the Flanker task (**Fig. S4D**). Participants viewed rows of five arrows or cartoon fish and were instructed to indicate the direction of the central stimulus while ignoring the flanking distractors. Each trial sequence included a fixation cross (800 ms), a response mapping cue (1,000-1,500 ms), a brief fixation (1,000 ms), and a stimulus array (100 ms). Up to 10,000 ms was allowed for response, recorded via button presses. Trials were categorized as congruent or incongruent based on directional alignment between flankers and targets. Flanker scores were generated through the NIH Cognitive Toolbox^15^. Specifically, task accuracy was the primary metric used to evaluate performance. For participants who achieved ≥80% accuracy, reaction time (RT) was also incorporated into the performance index. See Zelazo et al.^15^, for details of the calculation of Flanker performance.

Mental health was assessed using raw scores from the Child Behavior Checklist (CBCL)^16^, where higher scores indicate greater behavioral or emotional concerns. Consistent with the procedure used for the SDQ, the raw scores were standardized (z-scored) to facilitate interpretability and comparability across subscales. CBCL includes eight subscales—anxious/depressed, withdrawn/depressed, somatic complaints, social problems, thought problems, attention problems, rule-breaking behavior, and aggressive behavior. Among them, three subscales, including anxious/depressed, withdrawn/depressed, and somatic complaints, are commonly aggregated to form the internalizing problems broadband scale. Two other subscales, including rule-breaking and aggressive behavior, comprise the externalizing problems broadband scale. The remaining subscales (social problems, thought problems, and attention problems) do not contribute to these broadband scores but are reported individually. Previous studies have demonstrated strong correlations between CBCL and SDQ dimensions. Specifically, CBCL subscales for internalizing problems, social problems, externalizing problems, and attention problems correspond to the emotional symptoms, peer problems, conduct problems, and hyperactivity/inattention problem subscales of the SDQ, respectively^17,18^. Therefore, only these four CBCL-derived factors were retained for replication analyses.

We applied the same GAMLSS framework to model age-related changes in EF using flanker task performance from the ABCD. Among the candidate distributions, the generalized gamma (GG) distribution provided the best fit, as determined by the Bayesian Information Criterion (BIC) and model convergence (**Fig. S5D**). Age effects were modeled using B-spline functions with 4 degrees of freedom and a polynomial degree of 2, both selected based on BIC (**Fig. S6D**). Sex was included as a covariate in all distributional parameters. Model diagnostics indicated a good fit to the empirical data (**Fig. S7D** **& Fig. S8D**). Individual deviation scores were computed by comparing each participant’s performance to the estimated age- and sex-specific normative centiles.

To assess the relationship between EF deviations and mental health symptoms, we implemented two modeling approaches: (1) generalized additive mixed models (GAMMs), with age modeled nonlinearly using thin plate splines with a fixed basis dimension (*k* = 3); and (2) linear mixed models (LMMs), with age treated as a linear covariate. In both models, EF deviation z-scores, age and sex were included as fixed effects, and participant ID was modeled as a random intercept to account for within-subject dependencies across repeated measurements. The optimal model was selected from GAMM and LMM based on the Akaike Information Criterion.

GAMM:

$\text{CBCL z-scores \textasciitilde EF deviation z-score}\text{s}$ + s (Age, k=3, fx=TRUE) + Sex + (1|subID)

LMM:

$\text{CBCL z-scores \textasciitilde EF deviation z-score}\text{s}$ + Age + Sex + (1|subID)

We also used the selected model to assess whether the associations between EF deviation and mental health varied by age. To do this, we introduced interaction terms between age and EF deviation z-scores using the same modeling frameworks.

GAMM:

$\text{CBCL z-scores \textasciitilde s}\text{ }\text{(Age, by=EF deviation z-score}\text{s}\text{, k=3, }\text{fx}\text{=F}\text{ALSE}\text{) }$ +

s (Age, k=3, fx= $\text{F}\text{ALSE}$) + Sex + (1|subID)	(7)

LMM:

$\text{CBCL z-scores \textasciitilde EF deviation z-score}\text{s}$ * Age + Sex + (1|subID)	(8)

Consistent with our primary analyses, we first drew 10,000 samples from the parametric bootstrap distribution based on the covariance matrix of parameter estimates. For each draw, age-specific slopes between CBCL scores and EF deviations were computed at 1,000 evenly spaced age points across the entire age span. The resulting distributions of age-specific slopes were then compared with the overall linear relationships to identify developmental periods showing significantly stronger or weaker EF effects relative to the overall association (**Fig. 4D**).

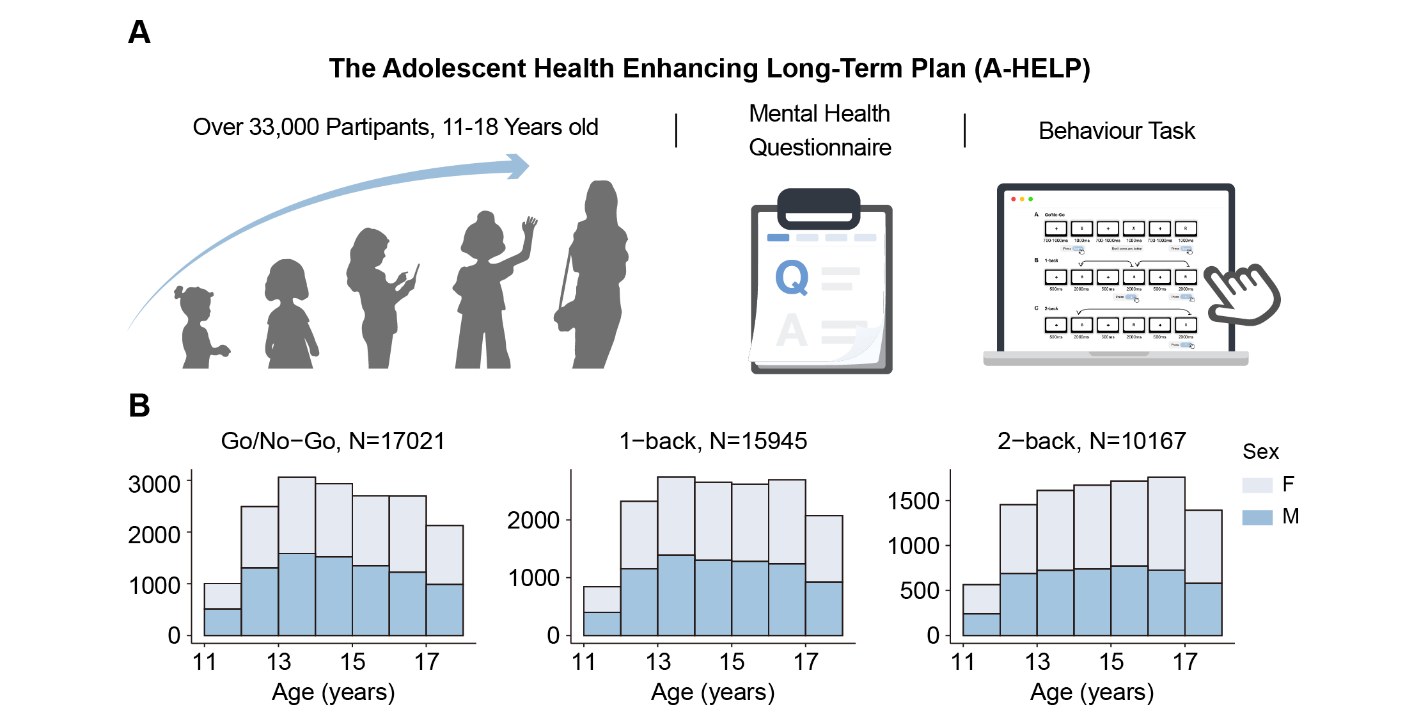
**Fig. S1.** Study design and final analytic sample. (**A**) Final sample sizes after applying all exclusion criteria were 33,622 Chinese adolescents (ages 11.00-18.00 years), who completed validated mental health questionnaires and standardized executive function tasks. (**B**) A total of 17,021 participants (8,485 males) were included for the Go/No-Go task, 15,945 (7,690 males) for the 1-back task, and 10,167 (4,468 males) for the 2-back task. Histograms show the age distributions of participants included in the analyses for each task (female in light blue, male in dark blue).

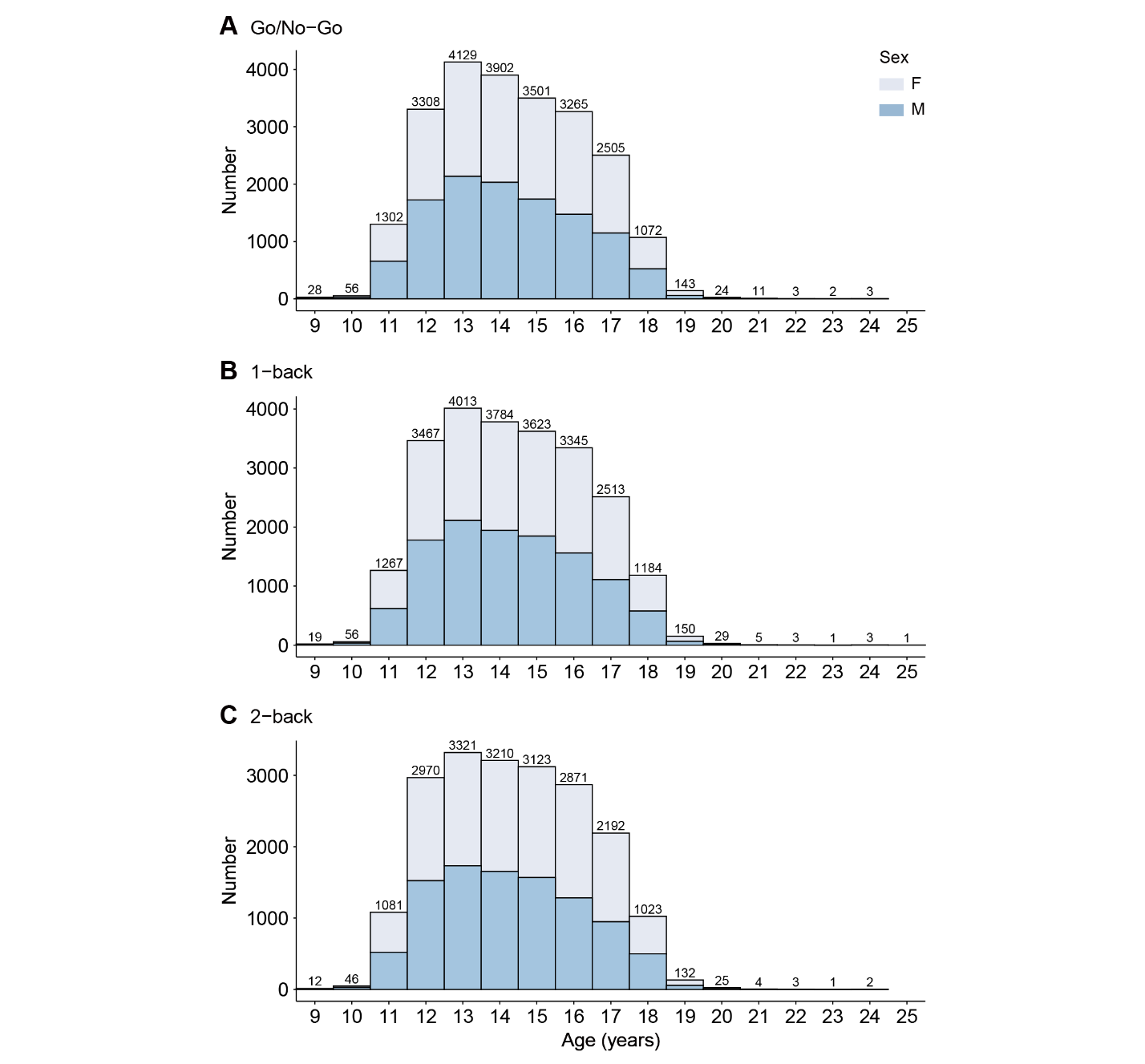

**Fig. S2. Age distribution of the raw included sample.** Age distribution of participants initially included in the three tasks (i.e., Go/No-Go, 1-back, 2-back), separated by sex. The displayed range (9-25 years) covers the vast majority of participants, while a small number of individuals younger than 9 or older than 25 were not shown due to their very limited counts. Participant numbers were sparse beyond 18 years, and subsequent analyses therefore primarily focused on the 11-18 years age range.

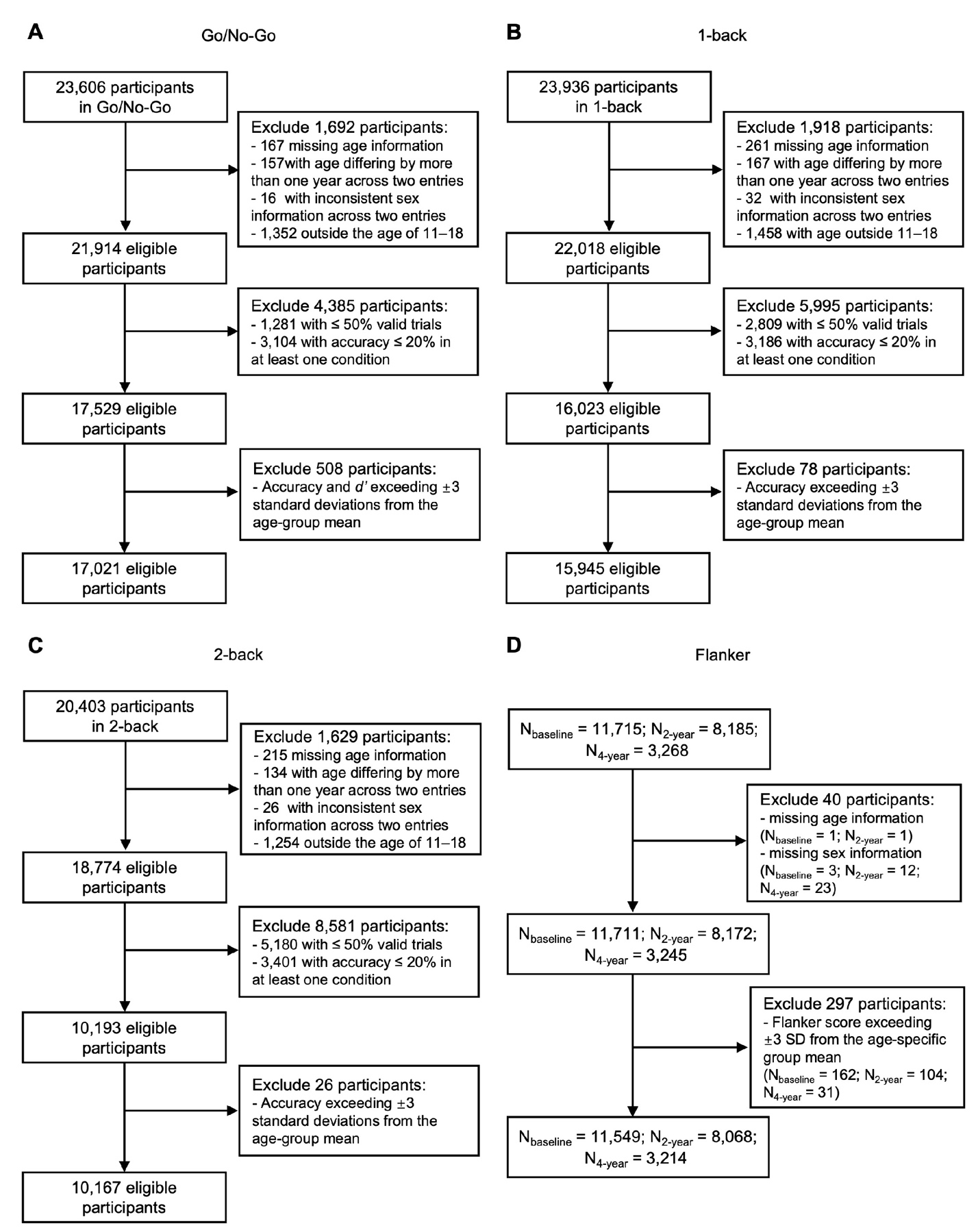

**Fig. S3. Flowchart of participant inclusion and exclusion for executive function tasks.** Panels A-C depict the sequential inclusion and exclusion steps for the Go/No-Go (**A**), 1-back (**B**), and 2-back (**C**) tasks from A-HELP cohort, starting from all participants with available data. Exclusion criteria included missing or inconsistent age information, age outside the target range of 11-18 years, insufficient valid trials (≤ 50%), and extreme outliers (> 3SD from age-specific means). The final eligible sample sizes were 17,301 for Go/No-Go, 15,963 for 1-back, and 10,173 for 2-back. Panel D shows the participant flow for the Flanker task from ABCD cohort, where exclusion was based on missing sex information and age-specific outlier detection (> 3 SD). Final eligible sample sizes were N_baseline_ = 11,549, N_2-year_ = 8,068, and N_4-year_ = 3,214.

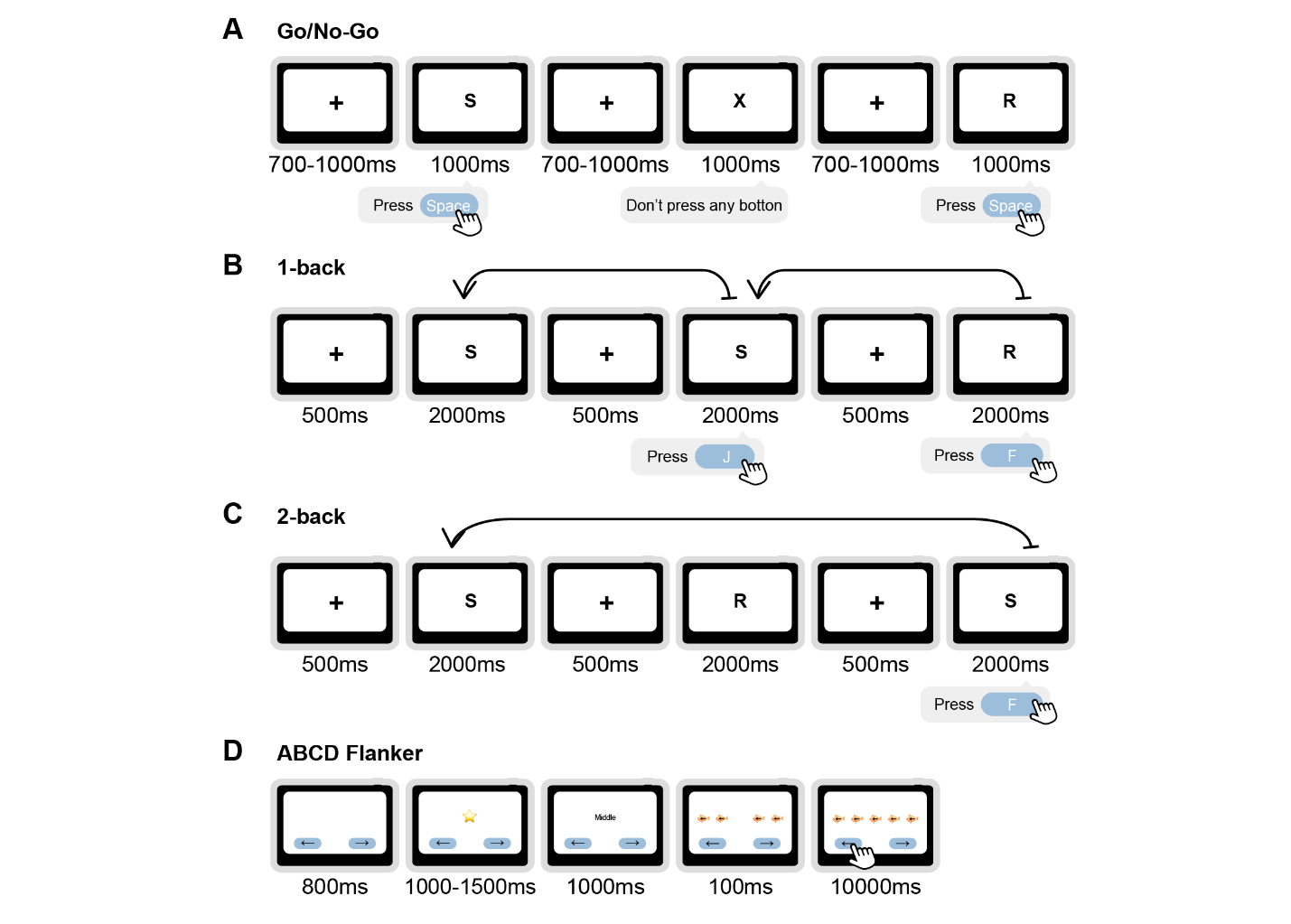

**Fig. S4.** **Schematic depiction of executive function tasks.** (**A**) Go/No-Go task. Participants were instructed to press the space bar in response to “Go” stimuli (any letter except “X”) and withhold responses to “No-Go” stimuli (“X”). Each trial consisted of a fixation cross (700-1,000 ms) and stimulus presentation (1,000 ms). (**B**) 1-back task. Participants were required to indicate whether the current stimulus matched the immediately preceding one. Each trial included a fixation cross (500 ms) and stimulus presentation (2,000 ms). (**C**) 2-back task. Participants were required to indicate whether the current stimulus matched the one presented two positions back in the sequence. Trial timing was identical to the 1-back task (fixation: 500 ms; stimulus: 2,000 ms). (**D**) Flanker task. Participants responded to the direction of a central target (fish or arrow) while ignoring surrounding flankers. Each trial began with a fixation cross (800 ms), followed by a cue screen indicating response mapping (1,000-1,500 ms), another fixation (1,000 ms), and finally the stimulus array (100 ms). Participants had up to 10 seconds to respond.

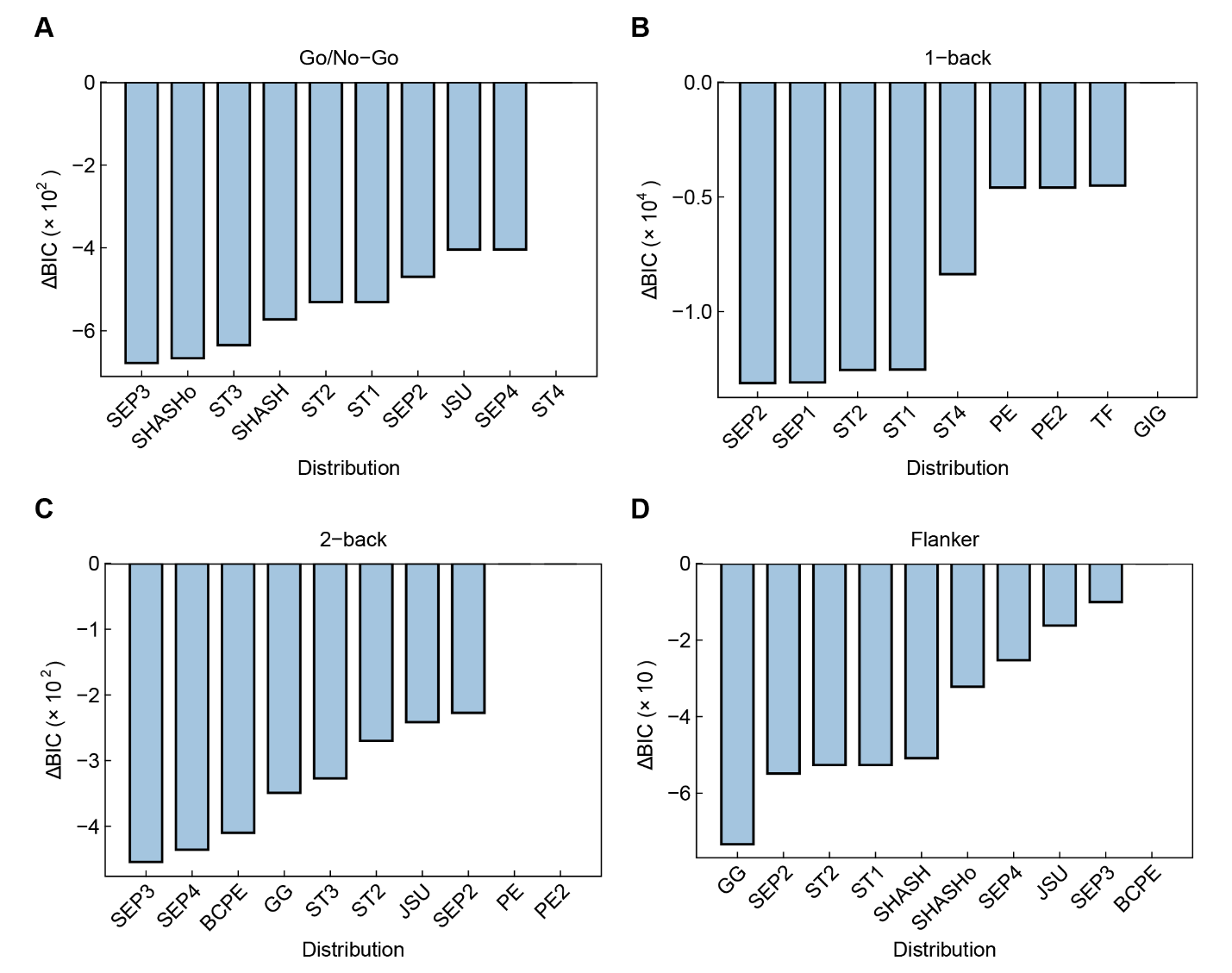

**Fig. S5.** **Bayesian information criterion (BIC) for candidate distribution families of executive function tasks.** For each task, the ten distributions with the lowest Bayesian Information Criterion values are shown, expressed as relative BIC (ΔBIC) after subtracting the BIC of the distribution with the largest value in that task. The largest BIC in each panel is therefore plotted as 0. The optimal distributions were SEP3 for Go/No-Go and 2-back, SEP2 for 1-back, and GG for Flanker. SEP2/3/4: Skew Exponential Power Types 2-4; ST1-4: Skew t Types 1-4; SHASH: Sinh-Arcsinh; SHASHo: Original Sinh-Arcsinh; JSU: Johnson’s SU; PE/PE2: Power Exponential; GG: Generalized Gamma; BCPE: Box-Cox Power Exponential; GIG: Generalized Inverse Gaussian; TF: t Family.

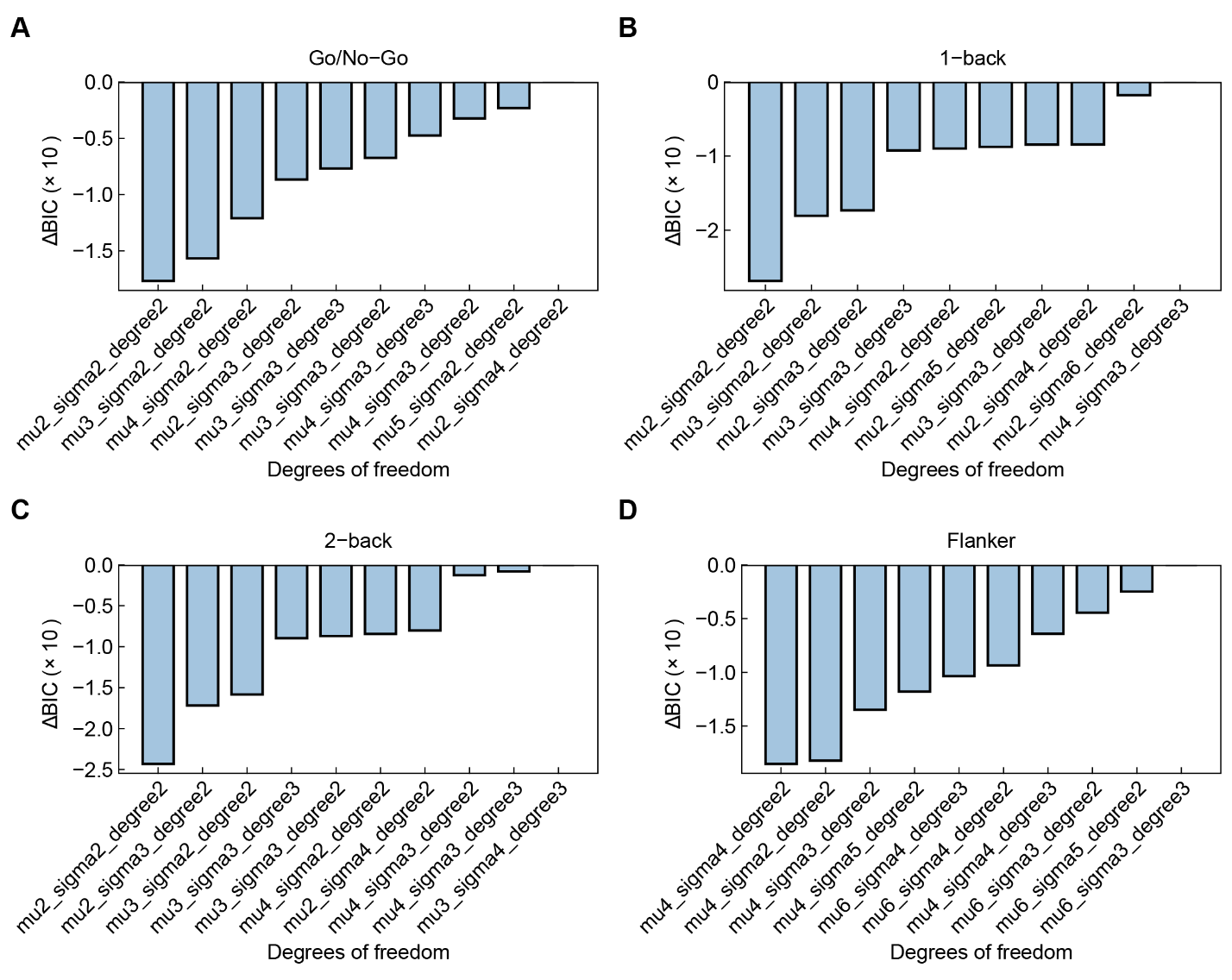

**Fig. S6. Bayesian information criterion (BIC) for parameter selection.** The optimal distribution families identified in Fig. S5 (SEP3 for Go/No-Go and 2-back, SEP2 for 1-back, and GG for Flanker) were further parameterized by varying the degrees of freedom (df) for the smooth terms of the location (*μ*) and scale (*σ*) parameters, as well as the polynomial degree of the B-spline basis for age. On the x-axis, labels indicate specific parameter settings: for example, “mu2” or “sigma4” indicate that the corresponding smooth term (*μ* and *σ*) was estimated with 2 or 4 degrees of freedom (df), respectively. The “degree” refers to the polynomial degree of the B-spline basis functions used to model smooth terms (degree = 2 indicates quadratic and degree = 3 indicates cubic). The y-axis shows relative BIC values, computed by subtracting the largest BIC among the compared specifications within each task. Thus, the best-fitting parameters for a given task are those with the smallest (i.e., most negative) relative BIC. Across tasks, the optimal specifications were: (**A**) Go/No-Go—*μ* = 2, *σ* = 2, degree = 2; (**B**) 1-back—*μ* = 2, *σ* = 2, degree = 2; (**C**) 2-back—*μ* = 2, *σ* = 2, degree = 2; and (**D**) Flanker—*μ* = 4, *σ* = 4, degree = 2.

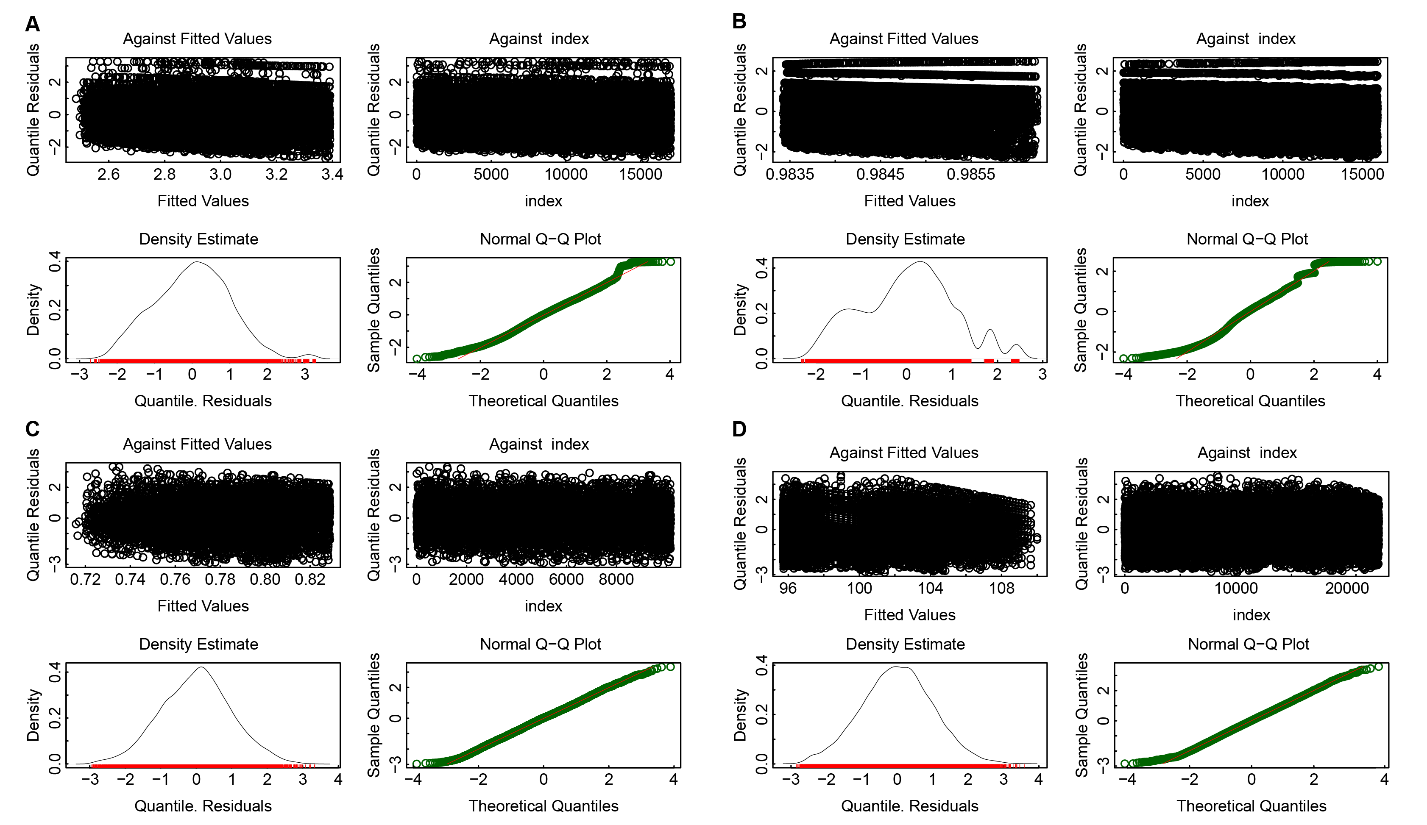

**Fig. S7. Residual distribution plots for the GAMLSS models.** Residual analyses are shown for (**A**) Go/No-Go, (**B**) 1-back, (**C**) 2-back, and (**D**) Flanker. Each panel includes four plots: residuals against fitted values, residuals against index, residual density estimates (black curve = observed residuals; red curve = reference normal distribution), and normal Q-Q plots. Overall, residuals for the four tasks approximate normality, supporting adequate model fit. Minor deviations are evident in the Go/No-Go and especially the 1-back tasks, where residual density and Q-Q plots reveal heavier tails and asymmetry relative to the theoretical normal distribution. In contrast, the 2-back and Flanker tasks show residual distributions closely aligned with normality, as indicated by density curves and near-linear Q-Q relationships.

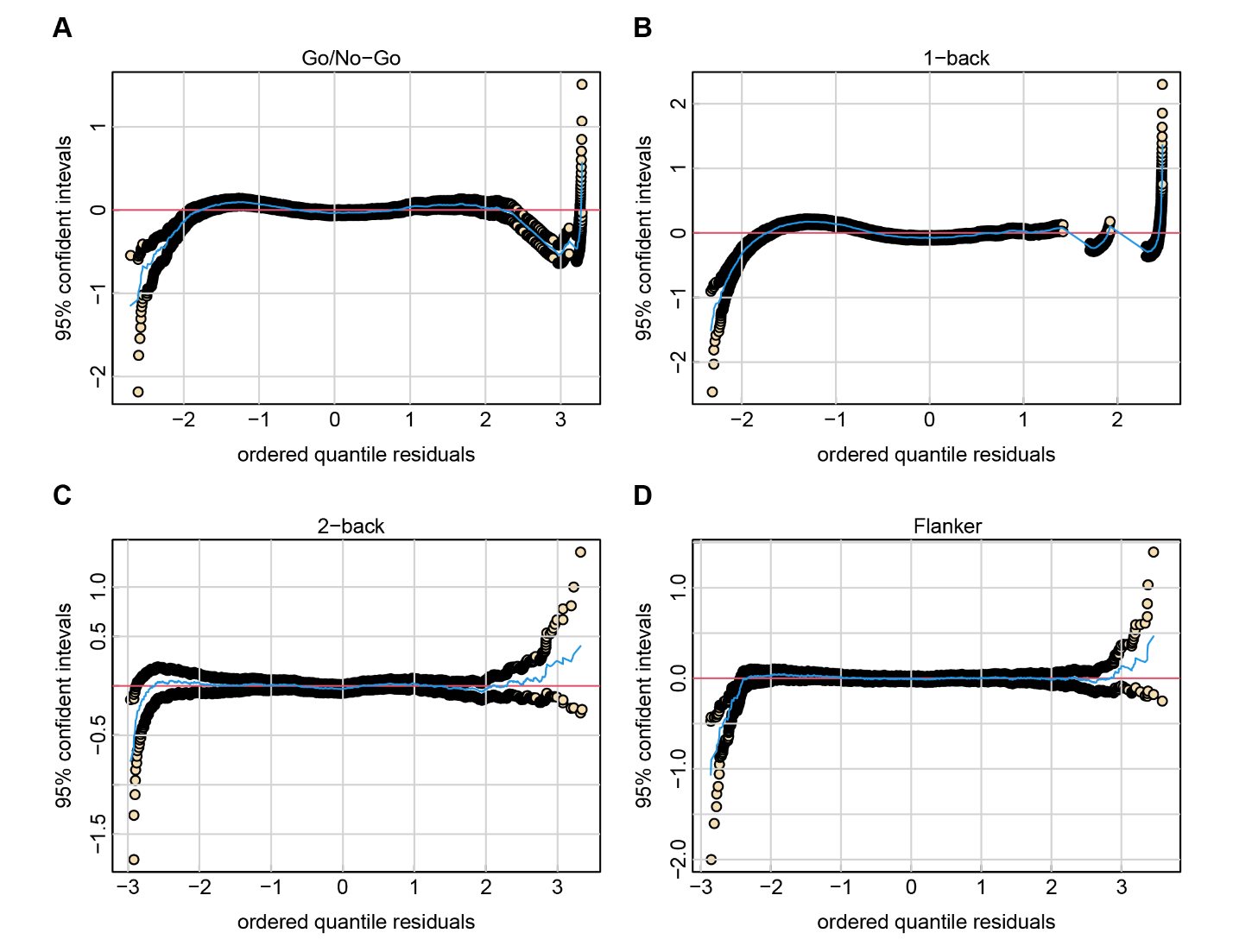

**Fig. S8. Detrended transformed Owen’s plots from the GAMLSS models.** Plots are shown for (**A**) Go/No-Go, (**B**) 1-back, (**C**) 2-back, and (**D**) Flanker. These diagnostic plots assess the adequacy of the model by comparing ordered quantile residuals against the expected theoretical distribution. The red horizontal line indicates zero, and the black curves represent 95% confidence intervals around the residuals. For all four tasks, the zero horizontal lines largely remained within the confidence intervals, suggesting approximate normality and acceptable model fit. Minor deviations at the distributional tails, particularly for the Go/No-Go and 1-back tasks, indicates some departures from normality but within a range commonly observed.

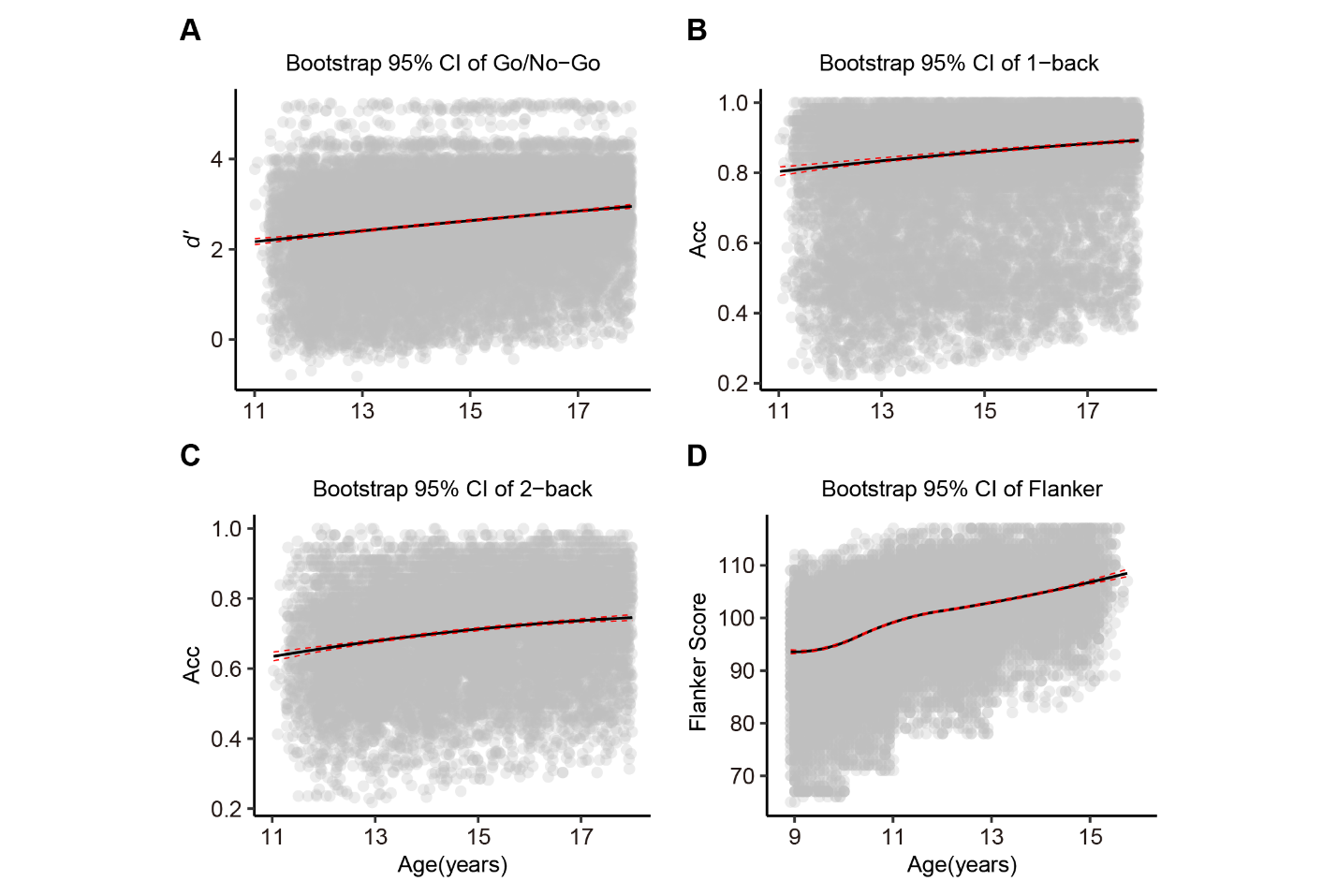

**Fig. S9. Bootstrap-derived 95% confidence intervals (CIs) for normative developmental trajectories.** The trajectories are shown for (**A**) Go/No-Go, (**B**) 1-back, (**C**) 2-back, and (**D**) Flanker tasks. Bootstrap analyses were performed using 10,000 iterations with replacement sampling, each followed by fitting GAMLSS models. The plots show the median (50^th^ percentile) developmental trajectories of task performance (black solid lines), with 95% confidence intervals (red dashed lines). Gray dots represent individual participants. Across four tasks, the bootstrap results demonstrate the robustness of the modeled developmental trajectories.

**Table S1. Demographic characteristics of participants in the A-HELP dataset.**

|  | Participants with  Go/No-Go task | Participants with  1-back task | Participants with 2-back task |
| --- | --- | --- | --- |
| N | 17021 | 15945 | 10167 |
| Age(years) (mean (SD)) | 14.69 (1.77) | 14.76 (1.77) | 14.81 (1.78) |
| Sex = Male (%) | 8485 (49.85) | 7690 (48.23) | 4468 (43.95) |
| Handedness (%) |  |  |  |
| Right-handed | 15526 (91.22) | 14704 (92.22) | 9438 (92.83) |
| Left-handed | 1495 (8.78) | 1241 (7.78) | 729 (7.17) |
| Ethnicity (%) |  |  |  |
| Han nationality | 16845 (98.97) | 15791 (99.03) | 10065 (99.00) |
| Others | 176 (1.03) | 154 (0.97) | 102 (1.00) |
| EF performance (mean (SD)) | 2.48 (1.04) | 0.80 (0.18) | 0.69 (0.15) |
| Mental health (mean (SD)) |  |  |  |
| Emotional problems | 2.25 (2.36) | 2.26 (2.36) | 2.29 (2.35) |
| Peer problems | 2.70 (1.68) | 2.67 (1.66) | 2.62 (1.65) |
| Conduct problems | 1.43 (1.67) | 1.36 (1.60) | 1.35 (1.56) |
| Hyperactivity | 3.49 (2.05) | 3.46 (2.06) | 3.45 (2.08) |
| Prosocial behavior | 6.52 (2.57) | 6.59 (2.53) | 6.66 (2.47) |

EF performance was assessed using *d’* for the Go/No-Go task and accuracy for the 1-back and 2-back tasks.

**Table S2. Demographic characteristics of participants with Flanker task in the ABCD cohort.**

|  | Baseline | 2-year follow up | 4-year follow up |
| --- | --- | --- | --- |
| N | 11549 | 8068 | 3214 |
| Age(years) (mean (SD)) | 9.91 (0.62) | 11.97 (0.65) | 14.08 (0.70) |
| Sex = Male (%) | 6010 (52.04) | 4254 (52.73) | 1677 (52.18) |
| Handedness (%) |  |  |  |
| Right-handed | 9187 (79.55) | 6288 (79.55) | 2631 (81.86) |
| Left-handed | 814 (7.05) | 541 (6.84) | 255 (7.93) |
| Mixed handed | 1548 (13.40) | 1075 (13.32) | 328 (10.21) |
| Race/ethnicity (%) |  |  |  |
| Hispanic | 2342 (20.28) | 1538 (19.06) | 654 (20.35) |
| Non-Hispanic Asian | 252 (2.18) | 170 (2.11) | 72 (2.24) |
| Non-Hispanic Black | 1675 (14.50) | 990 (12.27) | 339 (10.55) |
| Non-Hispanic White | 6066 (52.52) | 4542 (56.30) | 1818 (56.57) |
| Other | 1214 (10.50) | 828 (10.27) | 331 (10.30) |
| Flanker score (mean (SD)) | 94.44 (8.41) | 100.52 (6.98) | 104.61 (6.76) |
| Mental health (mean (SD)) |  |  |  |
| Internalizing symptoms | 5.03 (5.53) | 4.95 (5.62) | 5.27 (6.19) |
| Social problems | 1.60 (2.27) | 1.32 (2.07) | 1.04 (1.82) |
| Externalizing symptoms | 4.41 (5.80) | 3.92 (5.50) | 3.54 (5.17) |
| Attention problems | 2.95 (3.48) | 2.71 (3.31) | 2.57 (3.26) |

**Table S3. Coefficients of skewness, kurtosis and Filliben correlation for the four executive function measurements.**

|  | Go/No-Go | 1-back | 2-back | Flanker |
| --- | --- | --- | --- | --- |
| Skewness coefficient | 0.10 | 0.04 | -0.01 | 0.01 |
| Kurtosis coefficient | 2.92 | 2.57 | 2.94 | 2.87 |
| Filliben correlation coefficient | 0.99 | 0.99 | 1.00 | 1.00 |

The skewness coefficient reflects the degree of asymmetry in each distribution, with values close to zero indicating near symmetry. The kurtosis coefficient describes the “tailedness” of the distribution, with values near 3 suggesting approximate normality. The Filliben correlation coefficient provides a goodness-of-fit index for normal probability plots, where values near 1.00 indicate strong agreement with a normal distribution.
